# Atlas of plasma nuclear magnetic resonance biomarkers for health and disease in 118,461 individuals from the UK Biobank

**DOI:** 10.1101/2022.06.13.22276332

**Authors:** Heli Julkunen, Anna Cichońska, Mika Tiainen, Harri Koskela, Kristian Nybo, Valtteri Mäkelä, Jussi Nokso-Koivisto, Kati Kristiansson, Markus Perola, Veikko Salomaa, Pekka Jousilahti, Annamari Lundqvist, Antti J. Kangas, Pasi Soininen, Jeffrey C. Barrett, Peter Würtz

## Abstract

Blood lipids and metabolites are both markers of current health and indicators of risk for future disease. Here, we describe plasma nuclear magnetic resonance (NMR) biomarker data for 118,461 participants in the UK Biobank, an open resource for public health research with extensive clinical and genomic data. The biomarkers cover 249 measures of lipoprotein lipids, fatty acids, and small molecules such as amino acids, ketones, and glycolysis metabolites. We provide a systematic atlas of associations of these biomarkers to prevalence, incidence, and mortality of over 700 common diseases (biomarker-atlas.nightingale.cloud/). The results reveal a plethora of biomarker associations, including susceptibility to infectious diseases and risk for onset of various cancers, joint disorders, and mental health outcomes, indicating that abundant circulating lipids and metabolites are risk markers well beyond cardiometabolic diseases. Clustering analyses indicate similar biomarker association patterns across different types of diseases, such as liver diseases and polyneuropathies, suggesting latent systemic connectivity in the susceptibility to a diverse set of diseases. The release of NMR biomarker data at scale in the UK Biobank highlights the promise of metabolic profiling in large cohorts for public health research and translation.

## Introduction

UK Biobank is a prospective study of approximately 500,000 individuals who have volunteered to have their health information shared with scientists across the globe to advance public health research. This open resource is unique in its size and availability of extensive phenotypic and genomic data (Sudlow et al. 2015; Bycroft et al. 2018; Szustakowski et al. 2021). A selection of 30 routine blood biomarkers has previously been measured in the full cohort (Allen et al. 2021; Sinnott-Armstrong et al. 2021), but there is a unique opportunity to evaluate the public health relevance of a wider range of biomarkers and accelerating translation, as exemplified by genome-wide genotyping for population-based risk identification (Khera et al. 2018).

Here we describe detailed metabolic biomarkers quantified by nuclear magnetic resonance (NMR) spectroscopy of 118,461 baseline plasma samples, generated by Nightingale Health Plc (Figure 1a). The sample size is more than ten-fold larger than many of the largest metabolic profiling studies conducted to date (Würtz et al. 2017; Pietzner et al. 2021). The NMR biomarker panel comprises 249 measures of lipids and metabolites (Figure 1b). These data are now available to approved researchers through the UK Biobank Showcase for all aspects of public health research. Many studies are already using these novel biomarker data, spanning applications related to, for instance, risk prediction, causal analyses, genetic discovery and drug target validation (Julkunen et al. 2021; Smith et al. 2022; Borges et al. 2022; Liu et al. 2022; Bragg et al. 2021; Richardson et al. 2021; Nag et al. 2021; Bell et al. 2021; Fang et al. 2021; Ritchie et al. 2021). We present a comprehensive atlas of biomarker-disease associations (available at biomarker-atlas.nightingale.cloud), systematically examined across the 249 metabolic measures in relation to presence, future onset and mortality of over 700 disease outcomes (Figure 1c). We illustrate the use of the atlas for biomarker discovery and identification of connections between overall biomarker signatures for various diseases. We replicate the findings in over 30 000 individuals from five prospective cohorts in the Finnish Institute for Health and Welfare (THL) Biobank profiled using the same NMR platform. Our biomarker-disease atlas may serve as a starting point to move from biomarker discovery to more detailed analyses in biological and clinical context.

**Figure 1:**
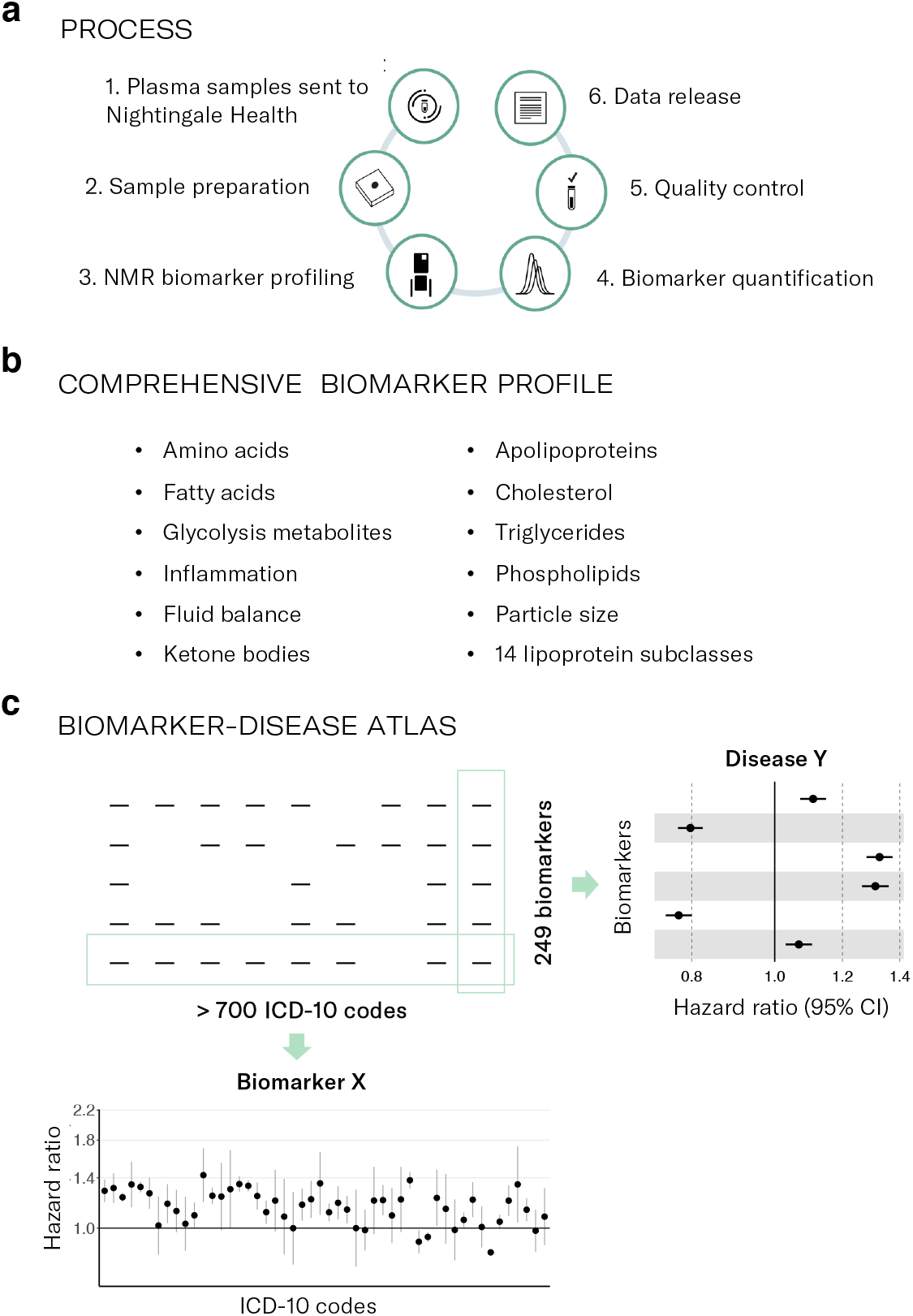
NMR biomarker data in the UK Biobank and atlas of disease associations. a) Process of the Nightingale Health-UK Biobank Initiative: 1) EDTA plasma samples from the baseline survey were prepared on 96-well plates and shipped to Nightingale Health laboratories in Finland, 2) Buffer was added and samples transferred to NMR tubes, 3) Samples were measured using six 500 MHz proton NMR spectrometers, 4) Automated spectral processing software was used to quantify 249 biomarker measures from each sample, 5) Quality control metrics based on blind duplicates and internal control samples were used to track consistency metrics throughout the project, 6) Biomarker data were cleaned, provided to UK Biobank and released to the research community. b) Overview of biomarker types included in the Nightingale Health NMR biomarker panel. c) Atlas of biomarker-disease associations published along with this study. The webtool allows to display the associations of all biomarkers versus prevalence, incidence and mortality of each disease endpoint, as well as show each biomarker versus all disease endpoints.

### Plasma biomarker profiling by NMR

We measured lipid and metabolite biomarkers from 118,461 baseline plasma samples using the Nightingale Health NMR platform (Figure 1a) (Soininen et al. 2015; Würtz et al. 2017; Julkunen et al. 2021). Table 1 shows characteristics of the participants with NMR biomarker data currently available in the UK Biobank. The EDTA plasma samples were picked randomly and are therefore representative of the 502,543 participants in the full cohort. Samples were generally drawn non-fasting, with an average of 4 hours since the last meal. The data release also contains biomarker measurements of approximately 4,000 repeat visit samples collected on average four years after the baseline, with approximately 1,500 participants having biomarker data from both baseline and the repeat-visit survey.

**Table 1.** Characteristics of the UK Biobank participants with plasma NMR biomarkers in the first data release from Nightingale Health.

|  | **Subset with**<br>**NMR**<br>**biomarkers,**<br>**baseline** | **Full cohort,**<br>**baseline** |
| --- | --- | --- |
| Number of participants | 118,461 | 502,543 |
| Age at blood sampling (median, [range]) | 58 [39-71] | 58 [37-73] |
| Females (%) | 54 | 54 |
| Body mass index (kg/m<sup>2</sup>), mean | 27.4 | 27.4 |
| Smoking prevalence (regular, occasional; %) | 7.9, 2.7 | 7.8, 2.7 |
| Fasting time, mean (h) | 3.8 | 3.8 |
| Self-reported cholesterol lowering medication use (%) | 18 | 17 |

The Nightingale Health NMR biomarker platform quantifies 249 metabolic measures from each sample in a single experimental assay, comprising 168 measures in absolute levels and 81 ratio measures (Figure 1b). The biomarkers include measures already routinely used in clinical practice, such as cholesterol, as well as many emerging biomarkers increasingly measured in cohorts, such omega-3 and other fatty acids (Würtz et al., 2017; Tikkanen et al. 2021). The panel of biomarkers is based on feasibility for accurate quantification in a high-throughput manner, and therefore mostly reflect molecules with high circulating concentration. Most of the biomarkers relate to lipoprotein metabolism, with the lipid concentrations and composition measured in 14 lipoprotein subclasses in terms of triglycerides, phospholipids, total cholesterol, cholesterol esters, and free cholesterol, and total lipid concentration within each subclass. The panel additionally includes the absolute concentration and relative balance of the most abundant plasma fatty acids, such as saturated fatty acids, and small molecules, like amino acids, and ketone bodies. Apolipoproteins B and A1, and two inflammatory protein measures, albumin and glycoprotein acetyls, are also measured, owing to their high abundance in plasma. Details of the NMR biomarker measurements in UK Biobank, including quality control, comparisons to other laboratory assays, and suggested approaches for data curation for epidemiological analyses are provided in Appendix 1.

### Atlas of biomarker-disease associations

The extensive electronic health records in the UK Biobank and the unprecedented sample size make it possible to study biomarker associations across the full spectrum of common diseases. We systematically computed the associations of the 249 NMR biomarkers with over 700 disease endpoints. Incident and mortality endpoints were defined by 3-character ICD-10 codes from nationwide hospital episode statistics and death records for diseases with at least 50 events occurring during 10 years after blood sampling. Prevalent endpoints were defined for diseases with over 50 events in the hospital records during approximately 25 years before the blood sampling. Details of the data pre-processing and statistical modelling are described in Methods. We collated the results in form of an online atlas of biomarker-disease associations available at biomarker-atlas.nightingale.cloud (Figure 1c). The webtool can display interactive forest plots for all biomarkers with prevalence, incidence, and mortality of each disease endpoint, as well as disease-wide association plots for each of the 249 biomarkers.

We observed a total of 33,764 individual biomarker associations to incident disease endpoints at *p* < 5e-5 (Methods). Similarly, for 648 prevalent disease endpoints and 77 causes of death, 26,035 and 3,055 significant associations were identified, respectively. These biomarker associations were not concentrated in cardiometabolic diseases, but spread across nearly all ICD-10 chapters. Examples include infectious diseases of both systemic and local character, certain cancers as well as mental and neurological disorders and musculoskeletal diseases. The magnitudes of biomarker associations for these diverse types of diseases were often similar to those of cardiovascular diseases. In the subsequent analyses in this paper, we focus on analyses of future onset of diseases from ICD-10 chapters A-N and the 37 biomarkers from the Nightingale Health NMR platform certified for diagnostic use.

### Biomarkers across the spectrum of diseases

Examining the NMR biomarkers across the spectrum of common diseases can provide insights into disease pathophysiology and specificity of the biomarkers. Figure 2a illustrates the span of diseases in different ICD-10 chapters associated with the 37 certified biomarkers. Many of the biomarkers exhibited associations across all types of diseases, with the exception of diseases of the eyes and the ears. For example, monounsaturated fatty acids relative to total fatty acids (MUFA%) was associated with almost 200 different disease endpoints spanning all ICD-10 chapters A-N. Also more established biomarkers such as omega-3% (i.e. concentration relative to total fatty acids) and routine cholesterol measures were associated with a wide spectrum of diseases. Glycolysis related metabolites and amino acids displayed fewer associations, but still spanning more than endocrine and circulatory diseases.

**Figure 2:**
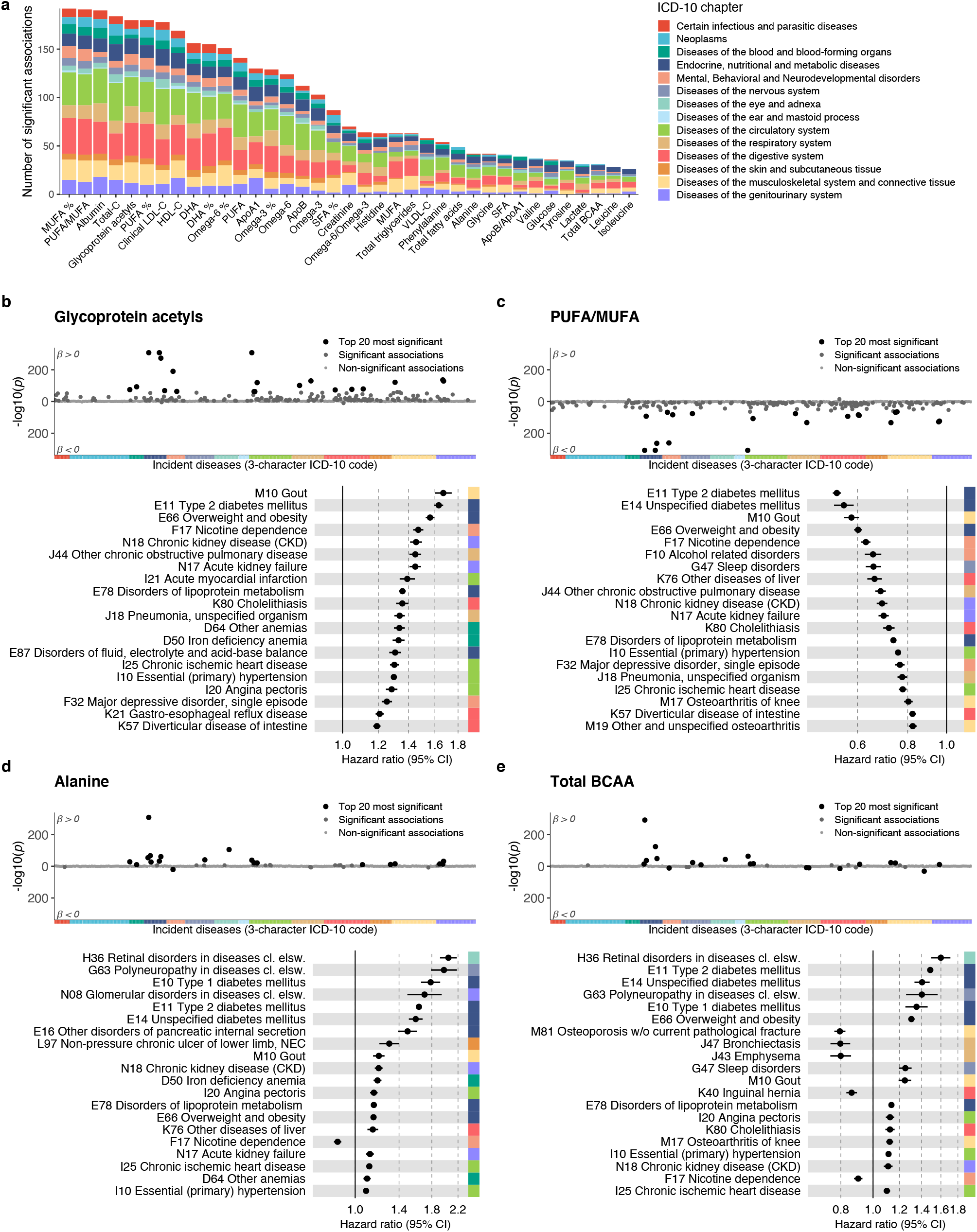
Biomarkers for future disease onset across a spectrum of diseases. a) Total number of incident disease associations by biomarker at statistical significance level *p* < 5e-5. The disease outcomes were 3-character ICD-10 codes with 50 or more events from chapters A-N, with a total of 556 diseases tested for association. The color coding indicates the proportion of associations coming from each ICD-10 chapter from A to N. b-e) Twenty most significant associations for four biomarkers: b) Glycoprotein acetyls, c) PUFA/MUFA, d) Alanine, and e) Branched-chain amino acids (BCAA). The top panel shows a disease-wide association plot of –log transformed p-values with incident diseases. Positive associations are displayed on the upper half of the plot and inverse associations on the bottom half. The color coding of the x-axis indicates the ICD-10 chapter, following the colors defined in panel a. The bottom panel highlights 20 of the most significant associations, arranged according to decreasing association magnitude. Hazard ratios and 95% confidence intervals are shown per SD-scaled biomarker concentrations. Similar disease-wide association plots for all 249 biomarkers across all endpoints analysed are available in the biomarker-disease atlas webtool.

Figure 2b-e shows the strongest incident disease associations in detail for four exemplar biomarkers; further examples are shown in Supplementary Figure 1. The inflammatory biomarker glycoprotein acetyls, also known as GlycA, was associated with the risk of 32% of the incident disease endpoints examined (*p* < 5e-5), with a median hazard ratio of 1.26 per 1-SD increment in the biomarker concentration. The most significant associations were observed for gout, type 2 diabetes, smoking dependence, kidney diseases, chronic obstructive pulmonary disorder, myocardial infarction, pneumonia and anemias. Figure 2c highlights the strongest disease associations for the ratio of polyunsaturated fatty acids to monounsaturated fatty acids (PUFA/MUFA), showing as widespread disease associations as for GlycA. Similar results were observed also for other fatty acid measures, such as omega-3% and omega-6% as well as MUFA% (Supplementary Figure 1a-c).

By contrast to this pattern of diverse associations, some biomarkers exhibited distinct disease specificity. For instance, the amino acid alanine was almost exclusively associated with the risk of diabetes and related complications (Figure 2d). Glycine and glutamine (Supplementary Figure 1d-e) were also associated with diabetes-related complications, but additionally with the risk of liver and kidney diseases, with lower plasma concentrations indicating higher disease risk. Glycine was also strongly associated with many circulatory disease endpoints, in line with the earlier suggested causal role of glycine levels in coronary heart disease (Wittemans et al. 2019). Most of the biomarkers had a consistent direction of associations across different diseases, but not all. For example, higher branched-chain amino acid levels were associated with a higher risk for many metabolic diseases but a lower risk for a range of other diseases such as lung diseases, hernia and smoking dependence (Figure 2e). A small number of biomarkers showed only weak magnitude of association across the spectrum of diseases, such as the ketone body 3-hydroxybutyrate (Supplementary Figure 1f).

Considered from the disease perspective, Figure 3 shows the biomarker association profiles for the incidence of six exemplar diseases. The results illustrates robust biomarker associations for incident hospitalisation for sleep disorders, depression, lung cancer and sepsis, with magnitudes of associations generally similar to those of myocardial infarction. The majority of the biomarkers associated exclusively in one direction of effect across these diseases and exhibited highly similar association patterns overall. An exception to this is osteoporosis, for which increased risk was characterized by decreased concentrations of branched-chain amino acids and triglycerides, and higher high-density lipoprotein (HDL) cholesterol and apolipoprotein A1 – in contrast to the other diseases in Figure 3.

**Figure 3:**
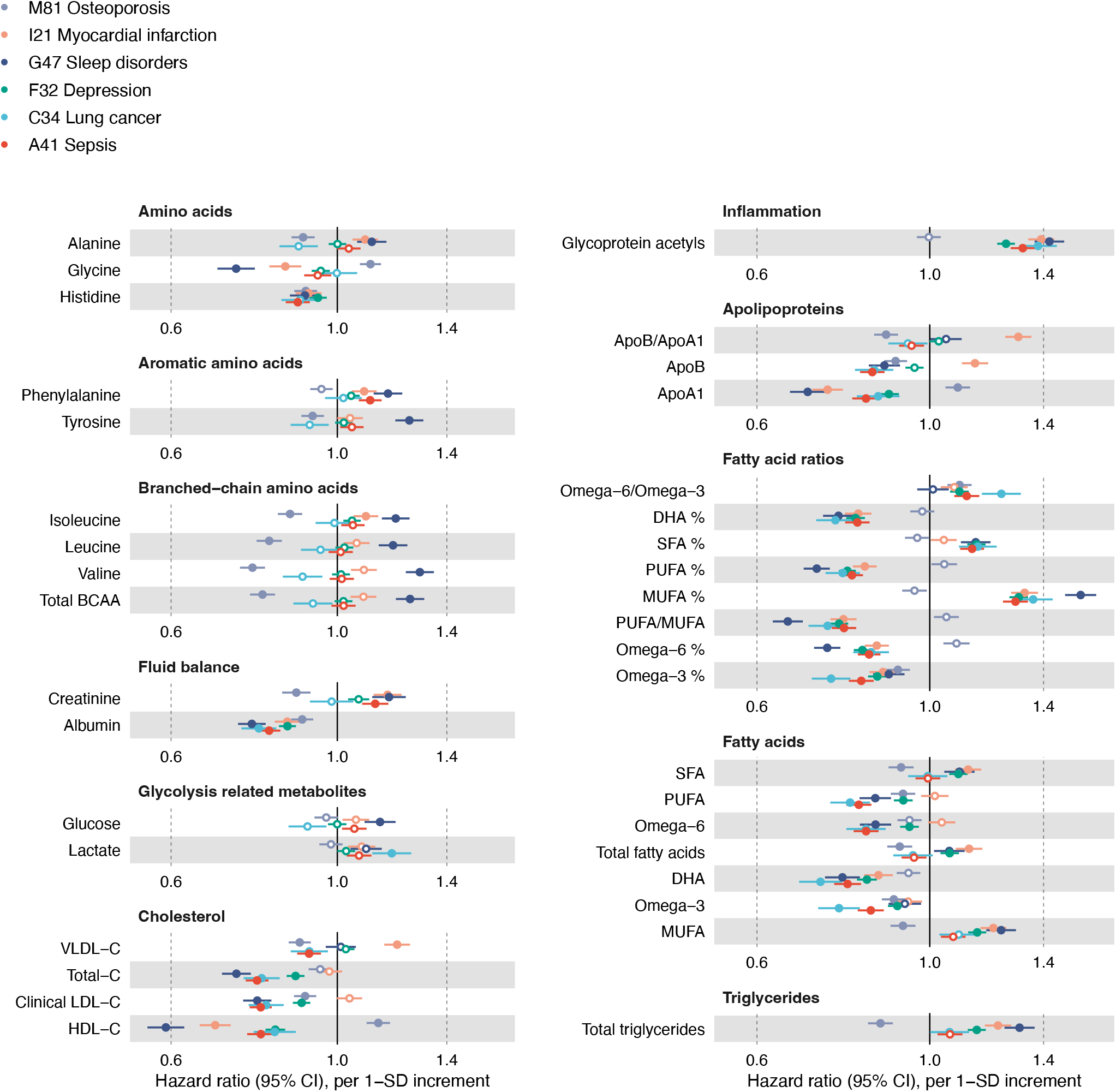
Biomarker profiles for various types of diseases. Hazard ratios of biomarkers with the incidence of six disease examples. Hazard ratios and 95% confidence intervals are shown per SD. The models were adjusted for age, sex and UK biobank assessment center, using age as the timescale of the Cox regression. Filled points indicate statistically significant associations (p < 5e-5), and hollow points non-significant ones. Similar forest plots for all 249 NMR biomarkers across all endpoints analysed are provided in the biomarker-disease atlas webtool.

### Shared biomarker signatures for different diseases

Comparing biomarker signatures between diseases may help to understand molecular differences between conditions with similar pathophysiology and identify novel connectivities (Pietzner et al. 2021; Holmes et al. 2018). Figure 4a shows examples of clustering of diseases according to their overall biomarker association patterns. On the vertical direction, biomarkers such as GlycA and MUFA% cluster together due their similarity in associations with many different types of diseases. Most amino acids cluster together, but glycine and histidine have deviating associations more similar to those of omega-6% and omega-3%, respectively. On the horizontal direction, the clustering analysis reveals both well-known connections between diseases and less anticipated similarities. For example, diabetes has a highly similar biomarker association patterns with several of its complications, including polyneuropathies and retinal disorders. Common diseases of an infectious origin, pneumonia and general bacterial infection, also cluster together in terms of their overall biomarker association patterns, as does COPD and lung cancer. Some of the less well-known connections include, for instance, liver diseases and polyneuropathies which had almost identical overall biomarker associations as further highlighted in Figure 4b.

**Figure 4:**
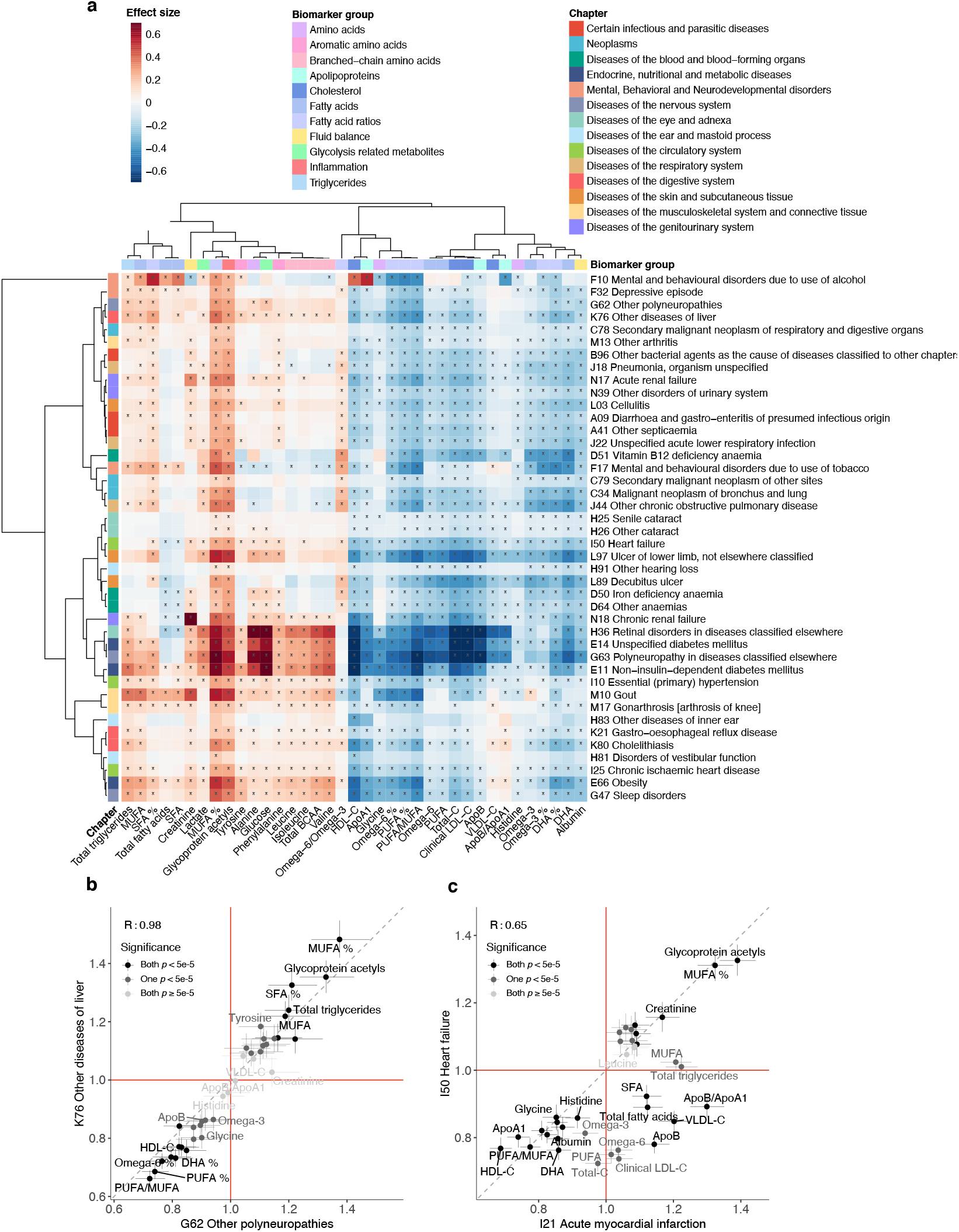
Clustering of diseases according to their biomarker signatures. a) Heatmap showing the clustering of biomarker association signatures across a diverse set of diseases. The diseases represent three diseases from each ICD-10 chapter from A to N selected based on the highest number of significant associations. The coloring indicates the association magnitudes in units of log(hazard ratio per SD). The dendrograms depict the similarity of the association patterns, computed using complete linkage clustering based on linear correlation between the association signatures. Significant associations with p-value < 5e-5 are marked with *. b-c) Examples of overall biomarker signatures compared for b) Other diseases of liver (K76) and Other polyneuropathies (G62), and c) Acute myocardial infarction (I21) and Heart failure (I50). The hazard ratios for each biomarker are shown as points with 95% confidence intervals indicated in vertical and horizontal error bars. The coloring of the points indicates the significance of the biomarker association for the pair of diseases. The red lines denote a hazard ratio of 1, and the grey line denotes the diagonal.

The biomarker signatures were similar for many diseases, but notable differences may still be observed for diseases of similar pathophysiological origin (Tikkanen et al. 2021). Figure 4c illustrates how acute myocardial infarction and hospitalisation for heart failure have many deviating biomarker associations even though these two endpoints are often combined for clinical trial analyses in the five-point major adverse cardiovascular event (MACE) definition. Supplementary Figures 2-4 further illustrate similarities and differences in the biomarker signatures for various other types of cardiovascular diseases. The biomarker association pattern differed for different types of myocardial infarction, angina, chronic ischemic heart disease, and different types of stroke. Even more pronounced differences were observed when comparing to heart failure and peripheral artery disease. In particular, many biomarker associations appeared to be stronger for other circulatory endpoints than for myocardial infarction and ischemic stroke. These results may suggest potential benefits for risk prediction separately for these types of cardiovascular events.

### Replication of biomarker signatures

Replication is essential in biomarker studies, no matter the sample size of the discovery analyses. We therefore sought to replicate the NMR biomarker associations in the UK Biobank in two ways: first by comparing the results to biomarkers measured by independent laboratory assays from the same UK Biobank samples, and second by analysing NMR biomarker data for over 30 000 participants from the Finnish Institute for Health and Welfare Biobank (THL biobank). Figure 5 shows the high concordance between disease associations for the eight biomarkers that have been measured by both NMR and clinical chemistry. The associations always have the same direction, and the hazard ratios are sometimes stronger for one assay and sometimes another, suggesting neither is systematically better at capturing disease association. Small deviations in the results may be because the plasma samples used for the NMR measurements were more affected by a known sample dilution issue than the corresponding serum samples used for clinical chemistry (Allen et al. 2020). The consistency between the NMR-based and clinical chemistry assays in absolute concentrations is further discussed in Appendix 1.

**Figure 5:**
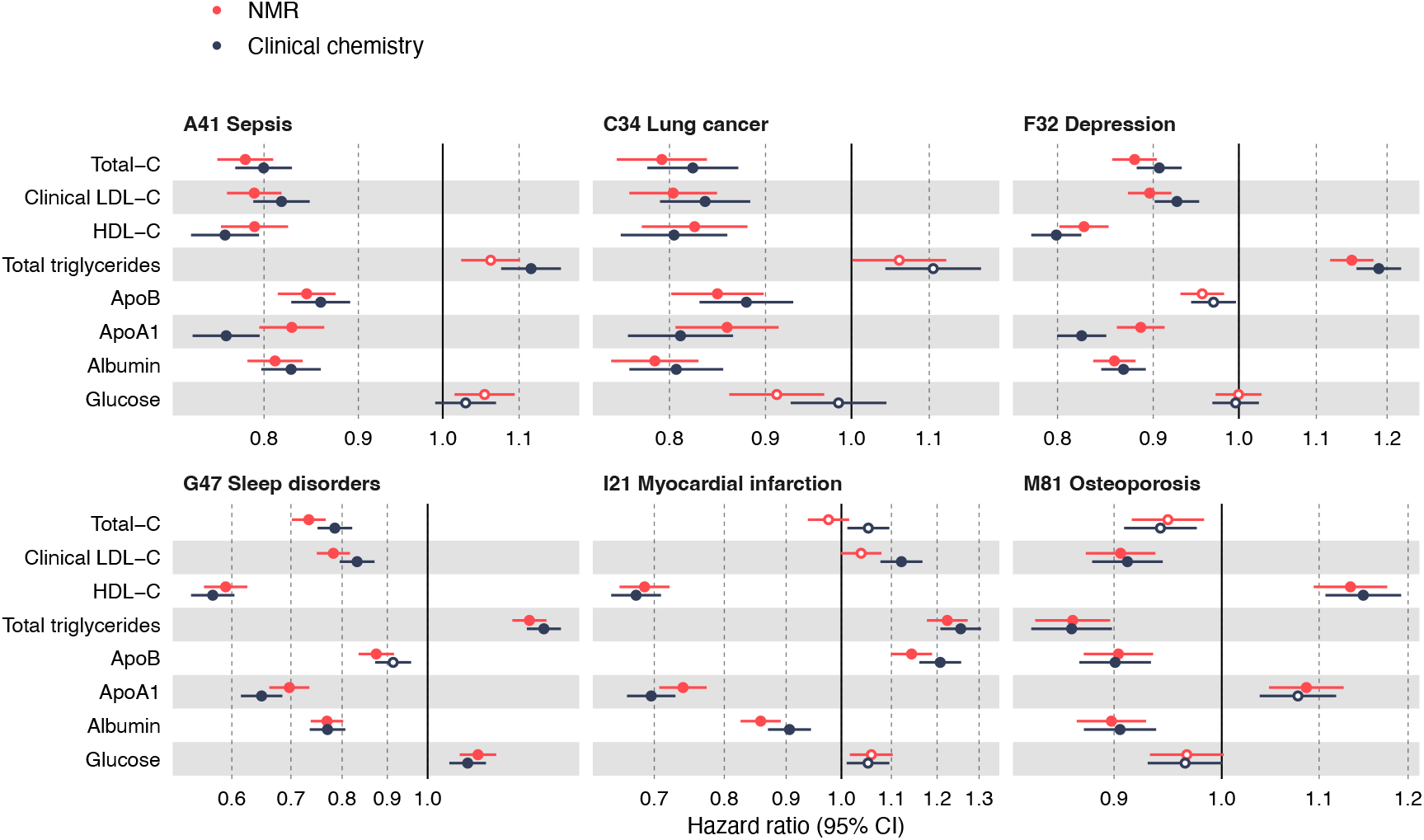
Comparison of NMR and clinical chemistry biomarker associations. Hazard ratios of biomarkers for which both NMR-based (in red) and clinical chemistry (in blue) measurements are available, against the incidence of six disease examples. Hazard ratios and 95% confidence intervals (CIs) are shown per SD. The models were adjusted for age, sex and UK biobank assessment center, using age as the timescale of the Cox regression. Filled points indicate statistically significant (p < 5e-5) associations, hollow points non-significant ones.

We note that low-density lipoprotein (LDL) cholesterol and apolipoprotein B displayed inverse associations across a wide range of diseases, i.e. higher concentration was associated with lower risk for disease incidence (Figure 5). This observation, which is surprising compared to existing literature on LDL as a risk factor, is seen in both the NMR and clinical chemistry measurements, indicating that it stems from characteristics of the UK Biobank study rather than any property of the NMR measurements. This observation was mainly explained by widespread use of lipid-lowering medications in the case of cardiovascular endpoints, since the inverse lipid associations were attenuated or flipped direction of effect when individuals on lipid-lowering medication were excluded (Supplementary Figure 5). Nonetheless, for most non-circulatory diseases, including five of the six disease examples shown in Figure 3, the LDL cholesterol associations remained inverse even after excluding individuals on lipid-lowering medication (Supplementary Figure 6) warranting further investigation in other cohorts.

We further replicated the associations observed in UK Biobank by a meta-analysis of five independent population-based cohorts from Finland measured using the same platform (Methods; clinical characteristics listed in Supplementary Table 1). Figure 6 illustrates the consistency of the biomarker association signatures against all-cause mortality and five available incident disease outcomes. Replication results for the remaining endpoints are shown in Supplementary Figure 7.

**Figure 6:**
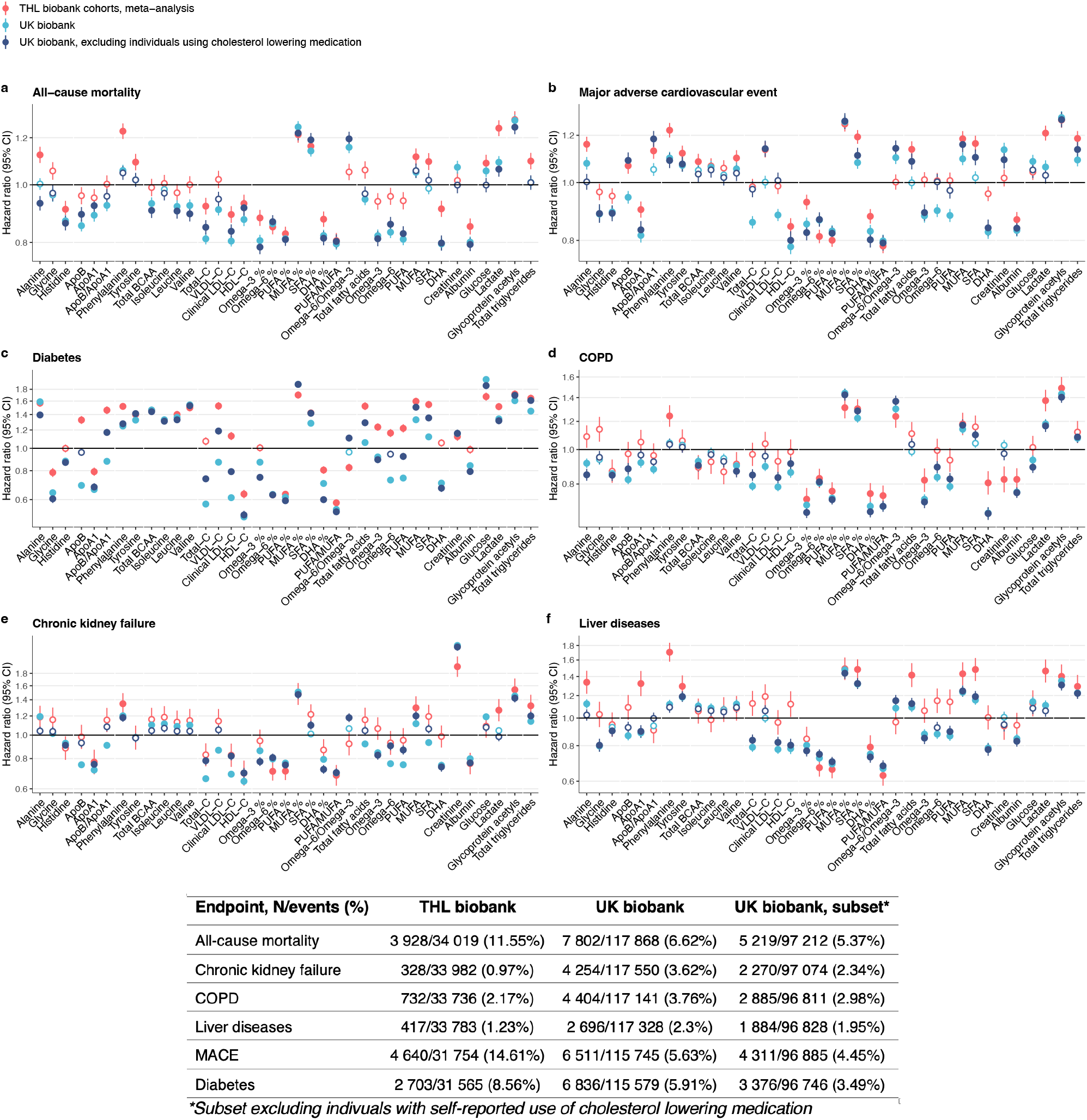
Replication of biomarker associations. Biomarker associations for six disease endpoints are shown for THL Biobank (red) and UK Biobank for the full study population (light blue) as well as for individuals without self-reported use of cholesterol-lowering medication (dark blue). Results from THL biobank were meta-analyzed for five prospective Finnish cohorts (FINRISK 1997, 2002, 2007, and 2012, and Health 2000). Hazard ratios and 95% confidence intervals (CIs) are shown per SD-scaled biomarker concentrations. Black horizontal line denotes a hazard ratio of 1. Event numbers for incident disease or mortality in the two biobanks are listed; ICD-10 codes used for compiling the composite endpoints are listed in Supplementary Table 2. The replication results are shown here for six endpoints available in THL biobank; results for all overlapping endpoints are shown in Supplementary Figure 7. Results are shown separately for each of the five Finnish cohorts in Supplementary Figure 8.

The biomarker associations were generally consistent in the two biobanks, especially for amino acids and other polar metabolites, fatty acid ratios and the two inflammatory protein measures. The greatest deviations were observed for aforementioned LDL-related biomarkers, which displayed strong inverse associations for diabetes and major adverse cardiovascular event in UK Biobank but flat or weakly positive in the Finnish cohorts. By excluding participants using cholesterol lowering medication in the UK Biobank, the associations generally became more consistent (Figure 6). However, many of the inverse associations for LDL cholesterol and related lipids also replicated in the Finnish cohorts, such as in the case of all-cause mortality and chronic kidney failure (Figure 6a and e). The results for absolute fatty acid concentrations deviated between the two study populations, whereas the results for fatty acid measures scaled relative to total fatty acids were highly concordant. This may suggest that such ratio measures are more easily transferrable across sampling approaches. The biomarker associations were consistent in each of the five Finnish cohorts, although there was a tendency for stronger hazard ratios for the cohort with shortest follow-up time (Supplementary Figure 8).

## Discussion

Detailed biomarker profiling is a key part of the promise of precision medicine initiatives to transform preventative healthcare. Blood biomarkers provide modifiable molecular measures which relate to future health outcomes and serve as intermediates between lifestyle factors and disease risk. This study describes the generation of NMR biomarker data by Nightingale Health in the UK Biobank, which is currently the world’s largest resource of metabolic biomarkers linked to health records. These data greatly extend the blood biomarker coverage in the UK Biobank and provide a wide span of molecular biomarkers not commonly measured in clinical practice, including amino acids, ketones and fatty acids. With over 118,000 plasma samples profiled in the UK Biobank, the addressable research questions extend vastly beyond biomarker discovery and the large sample size benefits, for example, causal analyses and risk prediction (Richardson et al. 2021; Fang et al. 2021; Julkunen et al. 2021; Bragg et al. 2022). Due to the streamlined data access policy in UK Biobank, the data release opens possibilities for the research community to use the entire epidemiological toolbox to study the NMR biomarkers in relation to public health.

The biomarkers in the Nightingale Health NMR platform are typically denoted ‘metabolic biomarkers’, and most prior studies on the data have focused on cardiometabolic diseases. Our analyses reveal that many of these biomarkers capture risk for many other diseases as well. This includes the future onset of diseases of the joints, bones, lungs, many different cancers as well as many mental disorders diseases and severe infectious diseases. These results explain earlier reports on strong associations of the NMR biomarkers with all-cause mortality (Deelen et al. 2019), since many of the biomarkers are associated broadly with leading causes of morbidity and mortality. Widespread associations across different diseases are known for inflammatory biomarkers such as GlycA (Ritchie et al. 2015; Kettunen et al. 2018), but it has not previously been shown for circulating fatty acids, amino acids or many detailed lipoprotein measures. For example, MUFA% was the biomarker associated across the highest number of endpoints and showed similar disease clustering as GlycA. Our results of widespread disease associations for many fatty acid ratios may suggest that these biomarkers should be considered as markers of systemic inflammation more so than of recent diet.

Plasma metabolites are increasingly understood to link to multimorbidities (Pietzner et al. 2021; Kettunen et al. 2018). This is strongly reinforced by our discovery of biomarker associations with the full spectrum of common diseases. We observed that a broad range of diseases with different pathophysiology were characterized by similar biomarker association profile. For example, severe infectious diseases had similar biomarker signatures to, for instance, chronic respiratory diseases as well as urinary and renal diseases. A potential explanation may be that many of the biomarkers reflect the innate immune system’s ability to respond. This would help to explain why many of the biomarkers were associated with susceptibility to severe infectious diseases, such as hospitalization and death from sepsis, fungal infections and pneumonia (Julkunen et al. 2021).

These observations illustrate how novel insights beyond individual diseases can be gained by studying overall biomarker signatures and numerous disease outcomes simultaneously. The genomic data in UK Biobank may help to elucidate causality of these results via Mendelian randomization (Fang et al. 2022; Borges et al. 2022).

Our biomarker-disease atlas published with this paper can be used to rapidly corroborate or refute many prior biomarker studies. For instance, we replicate the recent reports on higher branched-chain amino acid concentrations associated with lower risk for Alzheimer’s disease and dementia (Tynkkynen et al. 2018). The event numbers for these neurological diseases in UK Biobank alone are similar to those in the meta-analysed eight cohorts. The biomarker-disease atlas may also be used to put into question other reported biomarker discoveries, such as branched-chained amino acids in relation to risk for pancreatic cancer (Mayers et al. 2014): the association was essentially flat in UK Biobank despite a similar number of events. These examples illustrate how the biomarker-disease atlas may speed up research and serve as a starting point for analyses that yield deeper etiological insights and clinical context, much as widely available GWAS summary statistics transformed the interpretation of genetic studies. We note that the availability of the NMR biomarker data in UK Biobank does not diminish the relevance of having these data in smaller cohorts, both for replication and for complementary study designs. For example, the precise estimates of biomarker associations in UK Biobank can make analyses of smaller cohorts and trials more interpretable in relation to longitudinal sampling and intervention effects.

Metabolic profiling of all 500,000 baseline plasma samples in UK Biobank is underway. This will greatly expand the possibilities for studying rarer diseases and prediction of short-term risk, and also open possibilities for analyses focusing on individuals with prevalent disease and multi-morbidity trajectories. Coupled with the rich genomic data, clinical chemistry and proteomics measures, imaging, complete health-records, and other health related data that are continually added to the UK Biobank resource, the NMR biomarker data will enhance the possibilities for scientific discovery and is set to yield important findings for public health and clinical use. The data are available to approved researchers through similar access protocols as existing UK Biobank data (http://ukbiobank.ac.uk/).

## Methods

### UK Biobank cohort

The UK Biobank study was approved by the North West Multi-Centre Research Ethics Committee and all participants provided written informed consent. The study protocol is available online (https://www.ukbiobank.ac.uk). The biomarker profiling of plasma samples by NMR spectroscopy was approved under UK Biobank Project 30418.

The UK Biobank resource is a globally accessible biomedical database of half a million UK participants aged 40-69 years at baseline (Sudlow et al. 2015). A large variety of health information has been collected for each participant. For instance, the database includes questionnaire data on participant’s socio-economic and lifestyle factors, cognitive tests, imaging data, heart and lung function measures, body size and composition measures. Extensive genomic data is available, with genotyping array and exome-sequencing data available for all participants, and whole-genome sequencing under way (Bycroft et al. 2018).

The UK Biobank blood sample collection was undertaken at baseline in 22 local assessment centers across the UK between 2007 and 2010. The blood sample handling and storage protocol has been previously described (Elliot & Peakman. 2008). Prior to the measurement of the NMR biomarkers, 35 biomarkers have been measured from blood and urine samples (Allen et al. 2021, Sinnott-Armstrong et al. 2021).

### Plasma biomarker profiling by NMR

The main steps of the experimental procedures, including sample preparation and quality control, are described in Appendix 1. In order to serve as a reference for other studies using the data resource, this section also contains notes about plasma sample issues, comparisons to clinical chemistry and mass spectrometry as well as general recommendations for data processing in relation to epidemiological analyses.

### Disease outcome definitions

Prevalent, incident and mortality disease outcomes were derived from UK Hospital Episode Statistics data and national death registries. A diagnosis in hospital or death record formed the basis of the disease endpoint definition. Primary care records were not used. Disease endpoints were defined based on the first occurrence of 3-character ICD-10 code using the hospital inpatient and death register data (January 2021 update). To extend the follow-time prior to the introduction of ICD-10 in 1995, ICD-9 codes were mapped to the corresponding 3-character ICD-10 codes using general equivalence mappings from Center for Disease Control (https://ftp.cdc.gov/pub/Health_Statistics/NCHS/Publications/ICD10CM/2018/). A prevalent event was defined as an event that occurred before the date of participant’s baseline visit when a blood sample was collected. Individuals with corresponding prevalent event for each outcome were excluded from the analysis of incident disease, but not for analyses of mortality outcomes. The occurrence of both primary and secondary diagnoses codes was considered to form the endpoints. The follow-up of hospitalizations ended on November 30, 2020 in England, October 31, 2020 in Scotland, and February 28, 2018 in Wales. The follow-up of death registry ended on November 30, 2020. We omitted disease outcomes with fewer than 50 cases from the analyses. This led to a total of 648 prevalent, 717 incident and 77 mortality outcomes for the study population with NMR biomarker data available. For the examples highlighted in this paper, we focused on 556 incident disease outcomes from ICD-10 chapters A-N.

### Statistical methods

#### Biomarker association analyses across all endpoints

For the disease association analyses, biomarker values outside four interquartile ranges from median were considered outliers and excluded from the analyses. Furthermore, biomarker values were corrected for the NMR spectrometer used for the measurements by fitting a linear regression model with log1p-transformed concentrations as the outcome and spectrometer as the predictor. Scaled residuals from this regression were used as predictors in the association analyses.

We used Cox proportional hazard modelling to estimate associations between biomarkers and incident disease outcomes (hospitalization or death) across all endpoints with 50 or more events. The models were adjusted for sex and UK biobank assessment center, using age as the time scale of the Cox regression. Associations for each biomarker-disease pair were computed separately.

For biomarker association testing with prevalent diseases, we used logistic regression models adjusted for age, sex and assessment center. Hazard ratios and odds ratios are reported per SD increment in the log1p-transformed biomarker concentrations in order to allow comparison of association magnitudes for measures with different units and concentration range. Sex-specific diseases were conducted for 148 female diseases and 18 male diseases (Supplementary Table 3). These association analyses were performed in a subset containing only the specific sex, using the same approach without the inclusion of sex as a covariate.

In the biomarker-disease atlas, results are reported for all conducted analyses and the webtool allows to filter by a desired significance level. In this paper, we use a multiple testing-corrected significance level of 5×10^−5^ for reporting statistically significant associations, i.e. correcting for 1000 independent tests to account for both high correlation between the NMR biomarkers (∼50 independent tests; Würtz et al. 2017) and correlations between the disease endpoints analysed.

#### Clustering analyses

For clustering analyses, a dendrogram and heatmap was computed based on the association magnitudes of the 37 biomarkers with three diseases from each ICD-10 chapter from A to N. The diseases were selected based on the highest number of significant biomarker associations in each ICD-10 chapter. The 37 biomarkers selected are the ones clinically validated in the Nightingale Health NMR platform. Biomarkers are clustered in the dendrogram based on disease association profiles, and diseases are clustered based on biomarker profiles, using complete linkage clustering based on linear correlation between the association signatures.

#### Replication in additional cohorts

To replicate biomarker associations from the UK Biobank, we used data from five prospective population-based studies administered under the Finnish Institute for Health and Welfare (THL) Biobank: FINRISK 1997, FINRISK 2002, FINRISK 2007, FINRISK 2012 and Health 2000. Each cohort is an independent random sample drawn from people aged 25-98 (25–74 in FINRISK, 30 and over in Health 2000) in the Finnish population. The study participants are unique in each cohort. Baseline blood samples were collected for ∼85% of all participants enrolled. Venous blood was drawn non-fasting, but with recommended minimum of 4-h fast. Biomarker profiling by the Nightingale Health NMR platform was conducted from frozen serum samples for all participants during 2018 (Tikkanen et al. 2021). The cohort studies were approved by the Coordinating Ethical Committee of the Helsinki and Uusimaa Hospital District, Finland. Written informed consent was obtained from all participants.

Fourteen disease endpoints were used for replication analyses in THL Biobank, selected based on the outcome data made available to Nightingale Health Plc. The disease outcome definitions were pre-defined by THL Biobank based on a combination of national hospital and cause-of-death registries (Supplementary Table 2). The registry-based follow-up cover virtually all diseases leading to hospitalisation or death in Finland. Follow-up data for the present study were until the end of 2016. For the replication analyses, we defined similar endpoints in UK Biobank based on the ICD-10 codes listed in Supplementary Table 2.

The association analyses were for incident disease, so individuals with prevalent disease of the same endpoint were omitted. The hazard ratios were computed separately in each cohort using Cox regression adjusted for sex and using age as the time scale of the regression. Results from the individual cohorts were meta-analysed using inverse variance weighting. Similarly to the analyses in UK Biobank, hazard ratios are reported in SD-scaled units.

## Data Availability

The Nightingale Health NMR biomarker data have been released to the UK Biobank resource in spring 2021 (https://biobank.ndph.ox.ac.uk/showcase/label.cgi?id=220). The UK Biobank data are available for approved researchers through the UK Biobank data-access protocol.
Data from FINRISK and Health 2000 cohorts may be accessed through THL Biobank (https://thl.fi/en/web/thl-biobank).
As part of this publication, we provide access to all biomarker-disease summary statistics for academic use through an interactive webtool biomarker-atlas.nightingale.cloud.

https://thl.fi/en/web/thl-biobank

https://biomarker-atlas.nightingale.cloud

## Author Declarations

All relevant ethical guidelines have been followed; any necessary IRB and/or ethics committee approvals have been obtained and details of the IRB/oversight body are included in the manuscript.

All study participants provided informed consent. Ethical approval for UK Biobank was obtained from the North West Multi-Center Research Ethics Committee. The FINRISK and Health 2000 cohort studies were approved by the Coordinating Ethical Committee of the Helsinki and Uusimaa Hospital District, Finland.

## Acknowledgments

The authors are grateful to UK Biobank (Project #30418) and THL Biobank (project #BB2016_86) for access to data to undertake this study. The authors thank all biobank participants for their generous contribution to generating this resource for the scientific community.

## Data Availability

The Nightingale Health NMR biomarker data have been released to the UK Biobank resource in spring 2021 (https://biobank.ndph.ox.ac.uk/showcase/label.cgi?id=220). The UK Biobank data are available for approved researchers through the UK Biobank data-access protocol. Data from FINRISK and Health 2000 cohorts may be accessed through THL Biobank (https://thl.fi/en/web/thl-biobank).

As part of this publication, we provide access to all biomarker-disease summary statistics for academic use through an interactive webtool biomarker-atlas.nightingale.cloud.

## Contributions

HJ, AC, AJK, PS, JB, PW designed research; HJ and AC contributed to statistical analyses and interpretation of results; MT, HK, KN, VM, JN-K, PS, AJK contributed to biomarker measurements and quality control; MP, VS, PJ, AL, and KK contributed data or results for replication; HJ, AC, JB, and PW contributed to the interpretation of results and wrote the manuscript. All authors reviewed the manuscript.

