## Supplementary material for "Atlas of plasma nuclear magnetic resonance biomarkers for health and disease in 118,461 individuals from the UK Biobank"

**Supplementary Table 1. Baseline characteristics and event numbers in the five Finnish cohorts included in the replication analyses.**

| | **FINRISK 1997** | **FINRISK 2002** | **FINRISK 2007** | **FINRISK 2012** | **Health 2000** |
|---|---|---|---|---|---|
| Number of participants | 7580 | 7917 | 5966 | 5516 | 7100 |
| Age (median, [range]) | 49 [25-74] | 49 [25-74] | 52 [25-74] | 53 [25-74] | 53 [30-98] |
| Females (%) | 50 | 55 | 53 | 52 | 55 |
| Smoking prevalence (%) | 23.4 | 25.5 | 20.4 | 19.0 | 20.4 |
| Fasting time, mean (h) | 6.0 | 5.8 | 5.3 | 5.8 | 2.9 |
| Self-reported cholesterol lowering medication use (%) | 3.5 | 7.1 | 14.2 | 15.6 | 5.7 |

**Supplementary Table 2. Endpoint definitions for the replication analyses.**

| Pre-defined endpoint | ICD-10 codes used for endpoint definition in THL Biobank analyses | ICD-10 codes used for UK Biobank analyses |
|---|---|---|
| Major adverse cardiovascular event | I21-22, I50, I61, I63-64, I20.0, I11.0, I13.0, I13.2<br>Death records: I46, R96, R98 | I21-22, I50, I61, I63-64 |
| Diabetes | E10-E14* | E10-E14 |
| COPD | J43-J44 | J43-J44 |
| Chronic kidney failure | N18-N19 | N18-N19 |
| Liver diseases | K70-K77 | K70-K77 |
| Myocardial infarction | I21-22 | I21-22 |
| Heart failure | I50, I11.0, I13.0, I13.2 | I50 |
| Atrial fibrillation | I48 | I48 |
| Lung cancer | Lung cancer in cancer register | C34 |
| Fibrosis and cirrhosis of the liver | K74 | K74 |
| Rheumatoid disease | M05-M13, M32, M33, M45 | M05-M13, M32, M33, M45 |
| Heart disease mortality | I20-I25<br>Death records: I46, R96, R98 | I20-25 (death records) |
| Cancer mortality | Any cancer in cancer register | C00-C99 (death records) |

* Type 1 or type 2 diabetes; also national reimbursement records for anti-diabetic medication were use.

**Supplementary Table 3. Endpoints considered for sex-specific association analyses.**

| Sex | ICD-10 codes |
|---|---|
| Female | C50 Malignant neoplasm of breast |
| | C51-C58 Malignant neoplasms of female genital organs |
| | D05 Carcinoma in situ of breast |
| | D06 Carcinoma in situ of cervix uteri |
| | D25 Leiomyoma of uterus |
| | D26 Other benign neoplasms of uterus |
| | D27 Benign neoplasm of ovary |
| | D28 Benign neoplasm of other and unspecified female genital organs |
| | D39 Neoplasm of uncertain or unknown behaviour of female genital organs |
| | N60-N64 Disorders of breast |
| | N70-N77 Inflammatory diseases of female pelvic organs |
| | N80-N98 Noninflammatory disorders of female genital tract |
| | O00-O99 Pregnancy, childbirth and the puerperium |
| Male | N40-N51 Diseases of male genital organs |
| | C60-C63 Malignant neoplasms of male genital organs |
| | D29 Benign neoplasm of male genital organs |
| | D40 Neoplasm of uncertain or unknown behaviour of male genital organs |

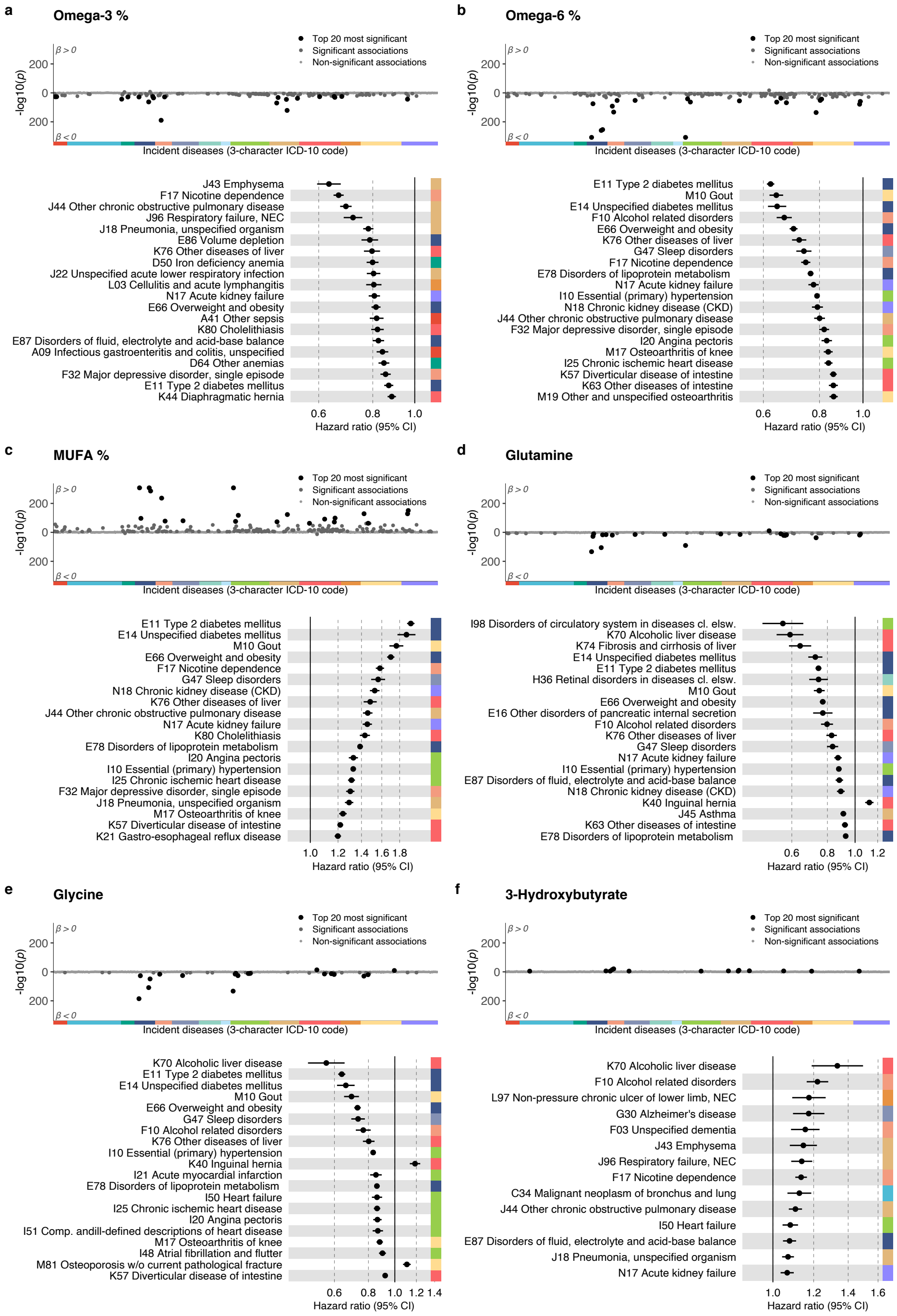


**Supplementary Figure 1: Examples of biomarkers for future disease onset.** Example of association discovery for six biomarkers: a) Omega-3%, b) Omega-6%, c) MUFA%, d) Glutamine, e) Glycine and f) 3-Hydroxybutyrate. The top panel shows a mirrored Manhattan-style plot of –log transformed p-values with the incidence of diseases with > 50 events across ICD-10 chapters from A to N. Positive associations are displayed on the upper half of the plot, inverse associations on the bottom half. The color coding of indicates distinct ICD-10 chapters, following the color coding in Figure 2. The bottom panel highlights 20 of the most significant associations, arranged according to a decreasing association magnitude. Hazard ratios and 95% confidence intervals (Cis) are shown per SD-scaled biomarker concentrations to facilitate comparison. Similar plots for all 249 biomarker measures across all endpoints analysed are available in the biomarker-disease atlas.

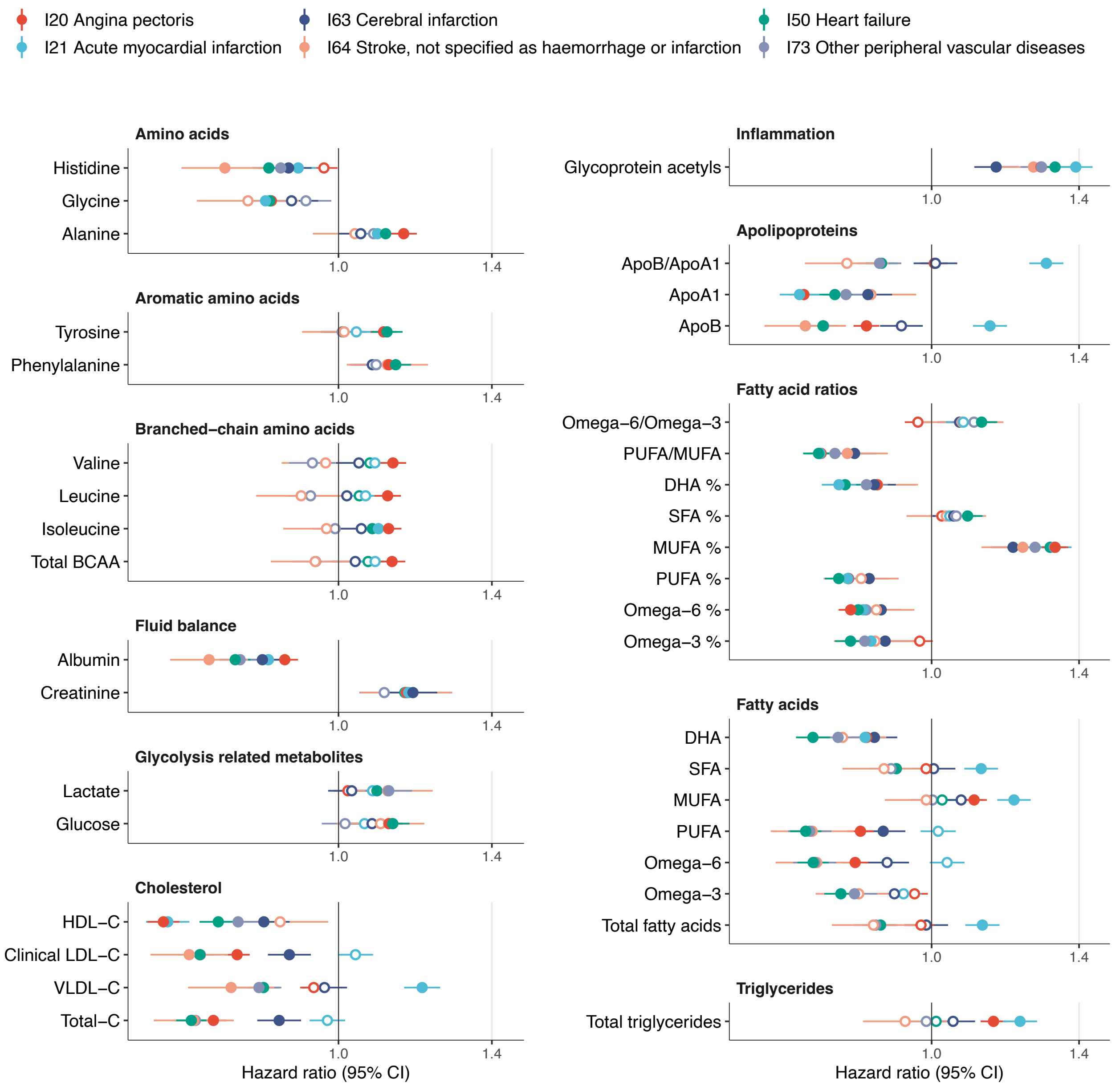


**Supplementary Figure 2. Biomarker profiles for incidence of cardiovascular diseases.**

Hazard ratios of biomarkers with the incidence of six cardiovascular disease endpoints. The biomarkers represent 37 biomarkers that are clinically validated in the Nightingale NMR platform. Hazard ratios and 95% confidence intervals (Cis) are shown per SD-scaled biomarker concentrations to facilitate comparison. The models were adjusted for age, sex and UK biobank assessment center, using age as the timescale of the Cox regression. Filled points indicate statistically significant ($p < 5e-5$) associations, hollow points non-significant ones.

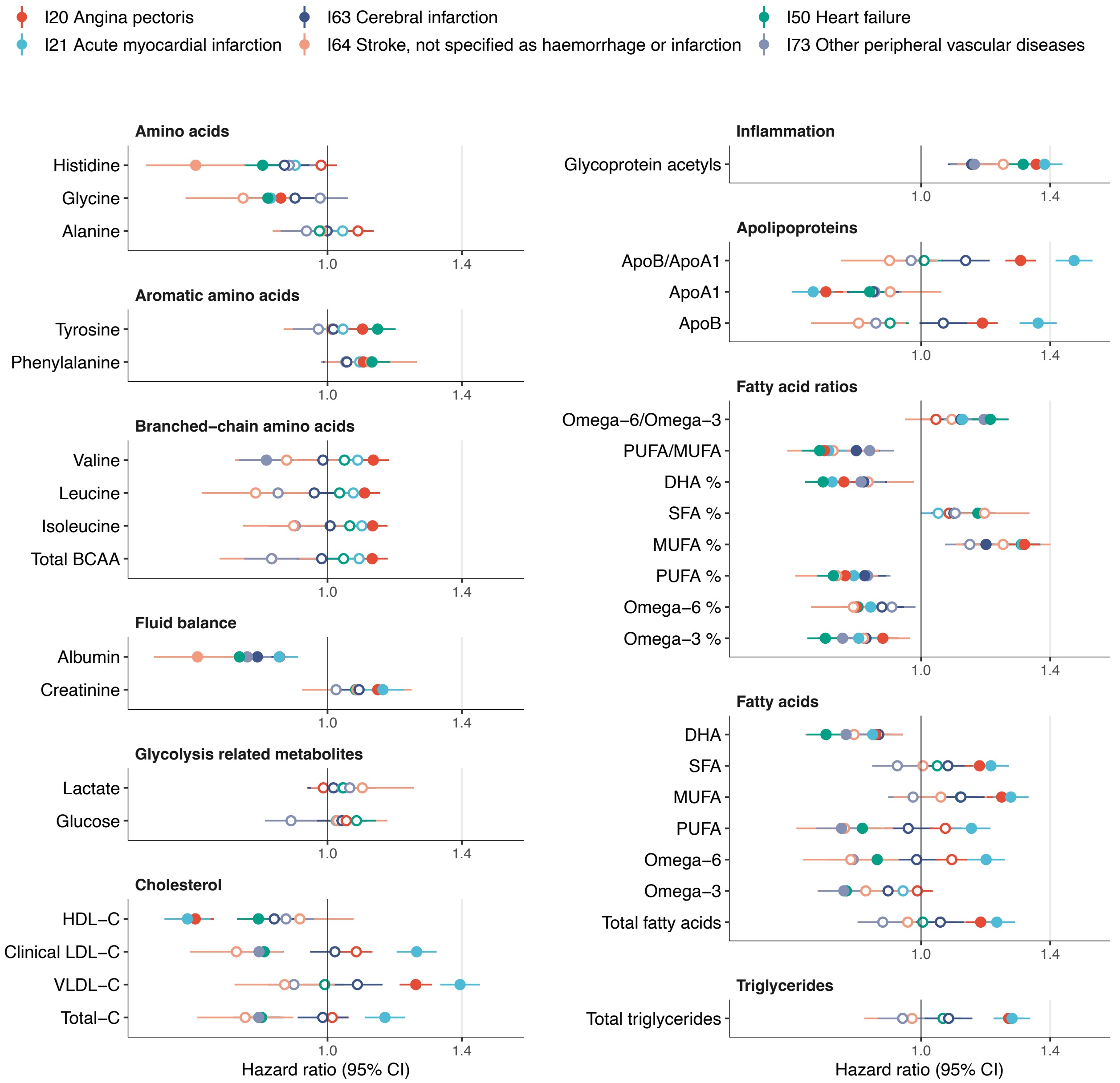


**Supplementary Figure 3. Biomarker profiles for incidence of cardiovascular diseases in a population free of cholesterol lowering medication.**
Hazard ratios of biomarkers with the incidence of six cardiovascular disease endpoints. The results are computed for a subset of UK biobank dataset excluding individuals with self-reported use of cholesterol lowering medication. The biomarkers represent 37 biomarkers that are clinically validated in the Nightingale NMR platform. Hazard ratios and 95% confidence intervals (Cis) are shown per SD-scaled biomarker concentrations to facilitate comparison. The models were adjusted for age, sex and UK biobank assessment center, using age as the timescale of the Cox regression. Filled points indicate statistically significant ($p < 5e-5$) associations, hollow points non-significant ones.

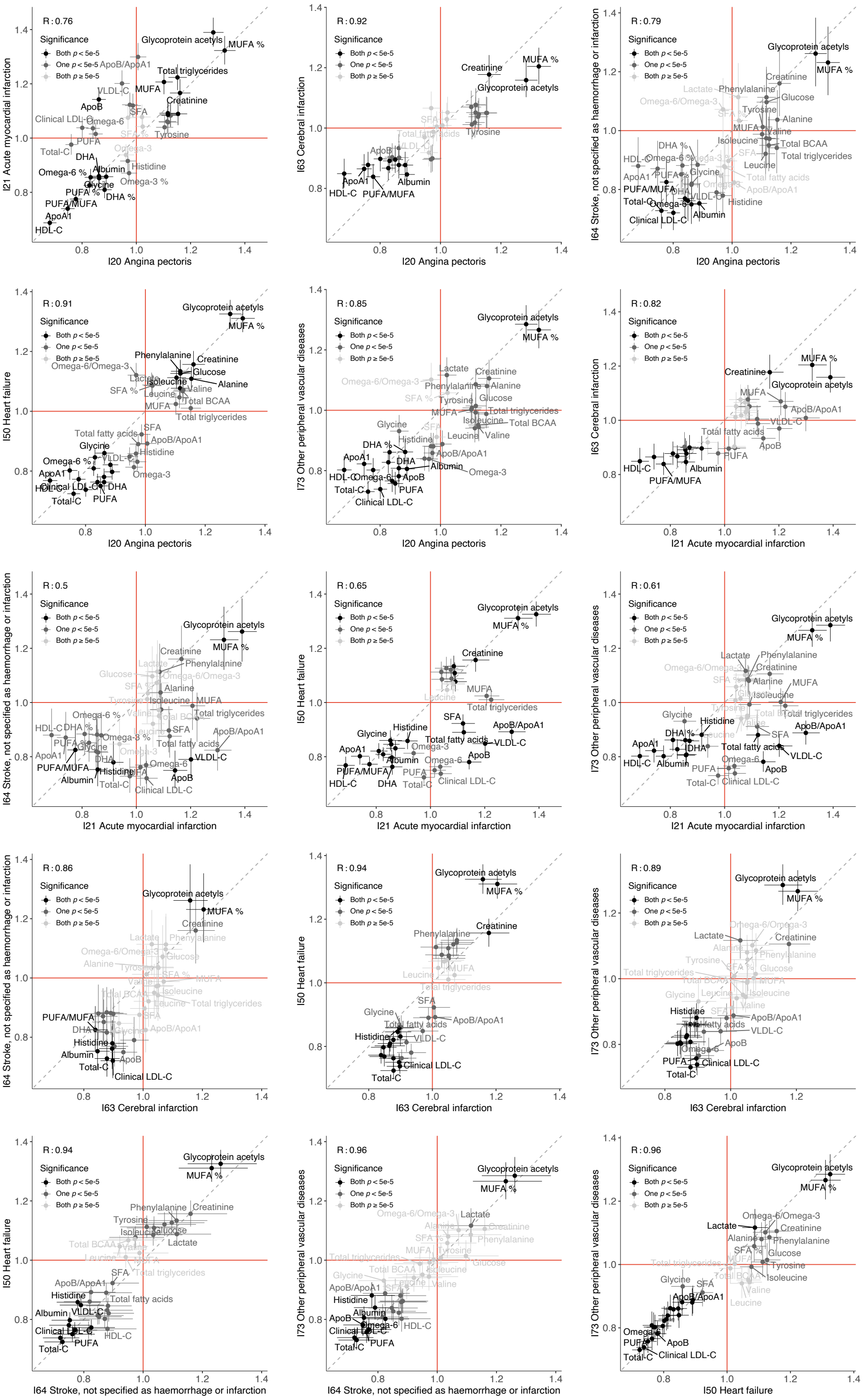


**Supplementary Figure 4. Correlation of biomarker association signatures for the incidence of cardiovascular diseases.**

Scatterplots of the association signatures between pairs of different cardiovascular disease endpoints. The hazard ratios for each biomarker (points) are given with 95% confidence intervals in vertical and horizontal error bars. The coloring of the points indicates the significance of the biomarker association for the pair of diseases. The red lines denote a hazard ratio of one, and the grey line denotes the diagonal.

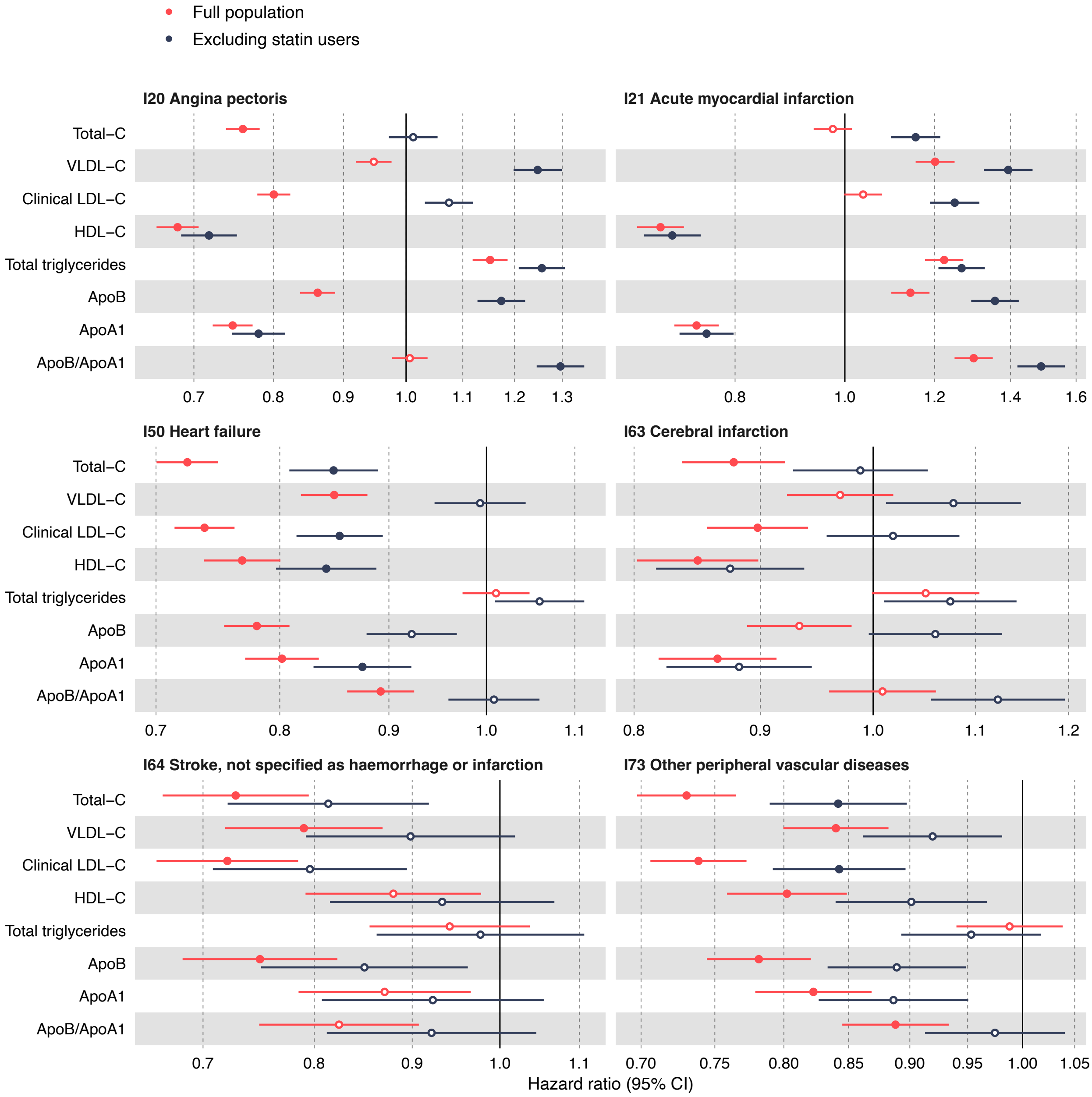


**Supplementary Figure 5. Influence of lipid lowering medication on associations with cardiovascular diseases.**
Hazard ratios for cholesterol and other lipid-related measures against the incidence of six cardiovascular diseases in the full UK biobank dataset (in red) and a subset excluding individuals with self-reported use of cholesterol lowering medication (in blue). Hazard ratios and 95% confidence intervals are shown per SD-scaled biomarker concentrations to facilitate comparison.

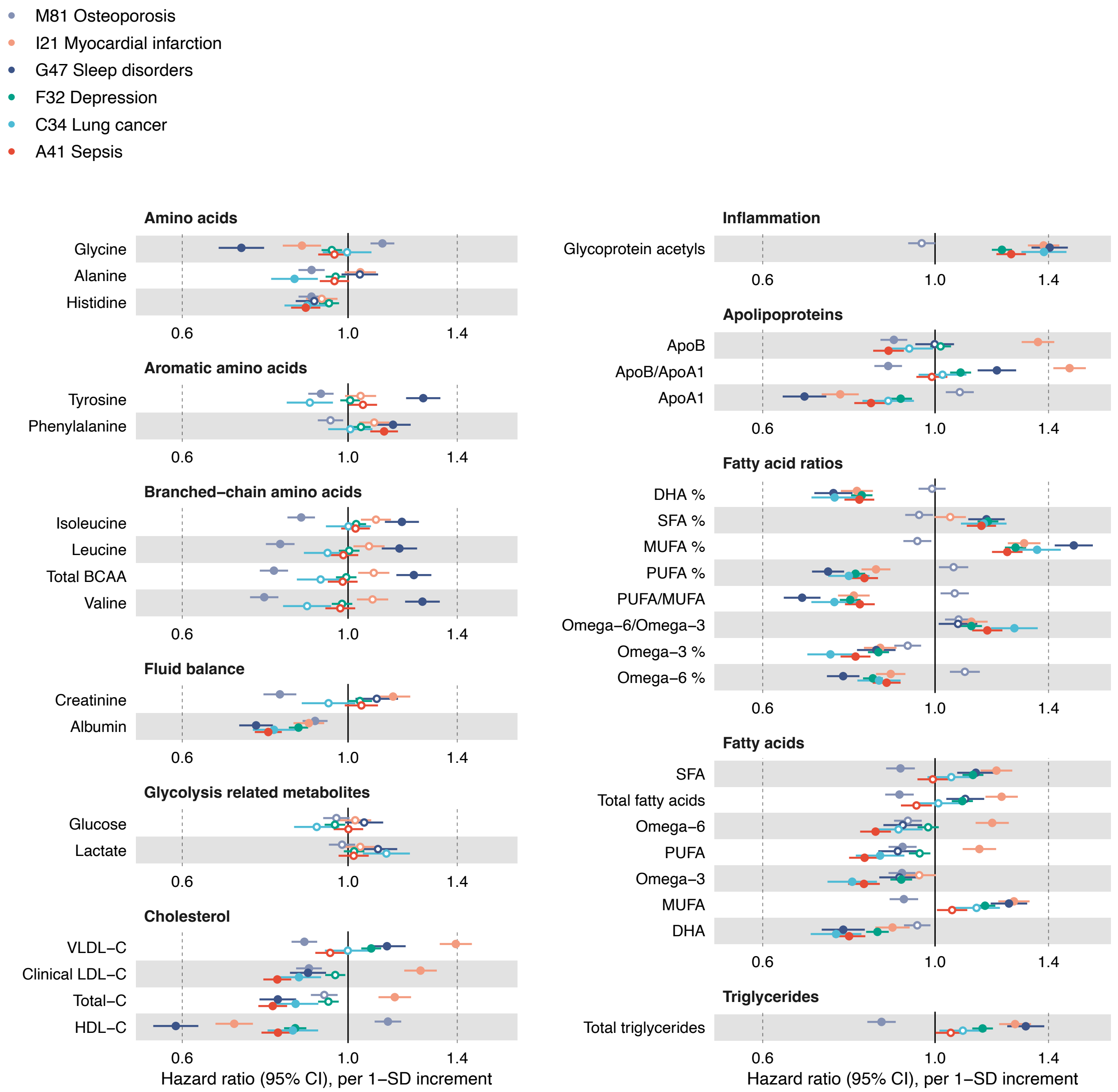


**Supplementary Figure 6. Biomarker profiles for various types of diseases in a population free of cholesterol lowering medication.**
Hazard ratios of biomarkers with the incidence of 6 selected example diseases from distinct ICD-10 chapters. The results are computed for a subset of UK biobank dataset excluding individuals with self-reported use of cholesterol lowering medication. The biomarkers represent 37 biomarkers that are clinically validated in the Nightingale NMR platform. Hazard ratios and 95% confidence intervals (Cis) are shown per SD-scaled biomarker concentrations to facilitate comparison. The models were adjusted for age, sex and UK biobank assessment center, using age as the timescale of the Cox regression. Filled points indicate statistically significant ($p < 5e-5$) associations, hollow points non-significant ones.

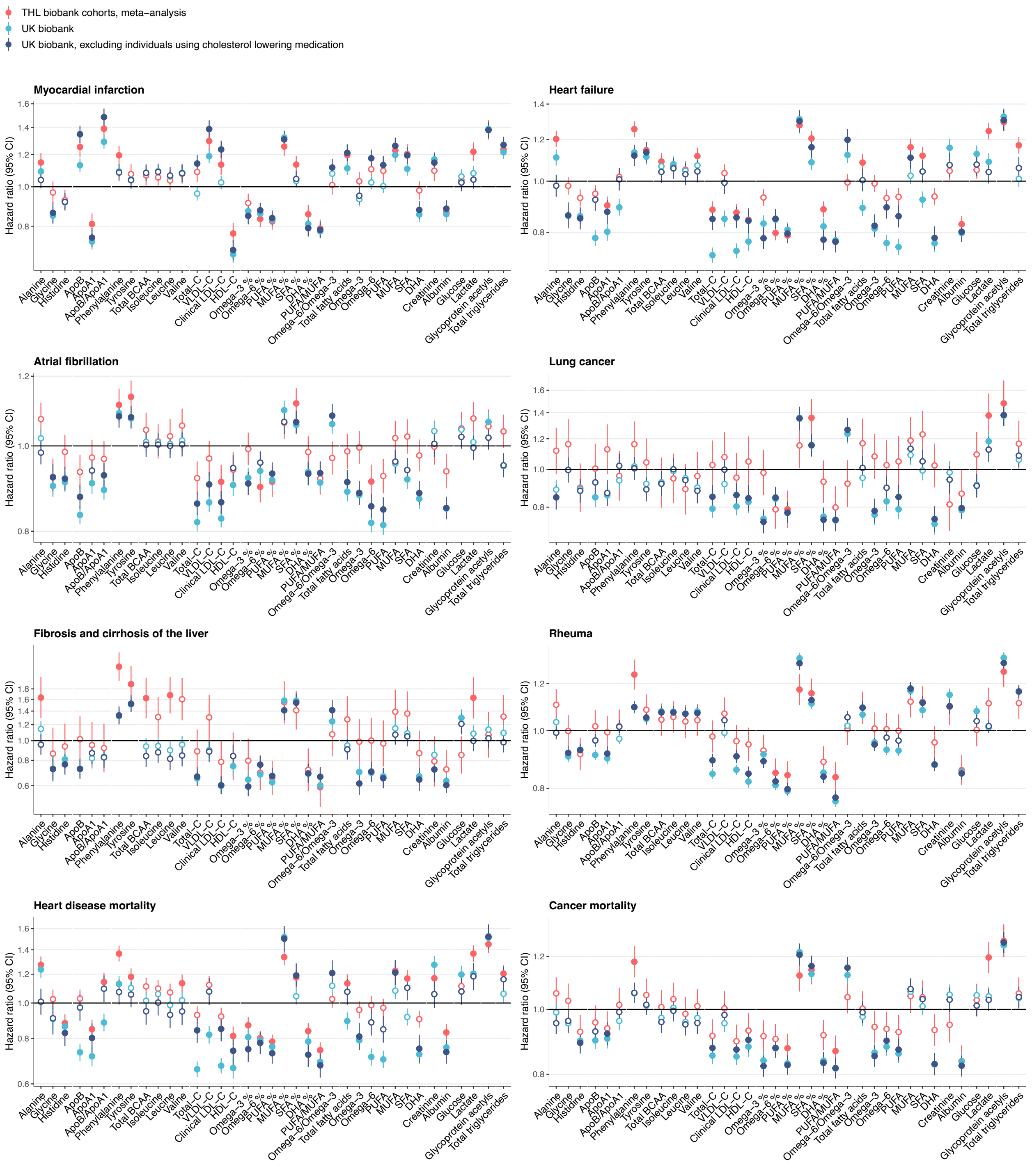


**Supplementary Figure 7. Replication of biomarker associations.**

Replication of biomarker associations across eight remaining endpoints in THL biobank; in red). Results from THL biobank represent meta-analyzed results for individuals from 5 prospective Finnish cohorts (FINRISK 1997, 2002, 2007, and 2012, and Health 2000). Results from UK Biobank are shown for the full population with NMR biomarker results available (light blue), as well as for a subset excluding individuals with self-reported use of cholesterol lowering medication (dark blue). Hazard ratios (HRs) and 95% confidence intervals (CIs) are shown per SD-scaled biomarker concentrations to facilitate comparison across the biomarkers. The black horizontal line denotes a hazard ratio of 1.

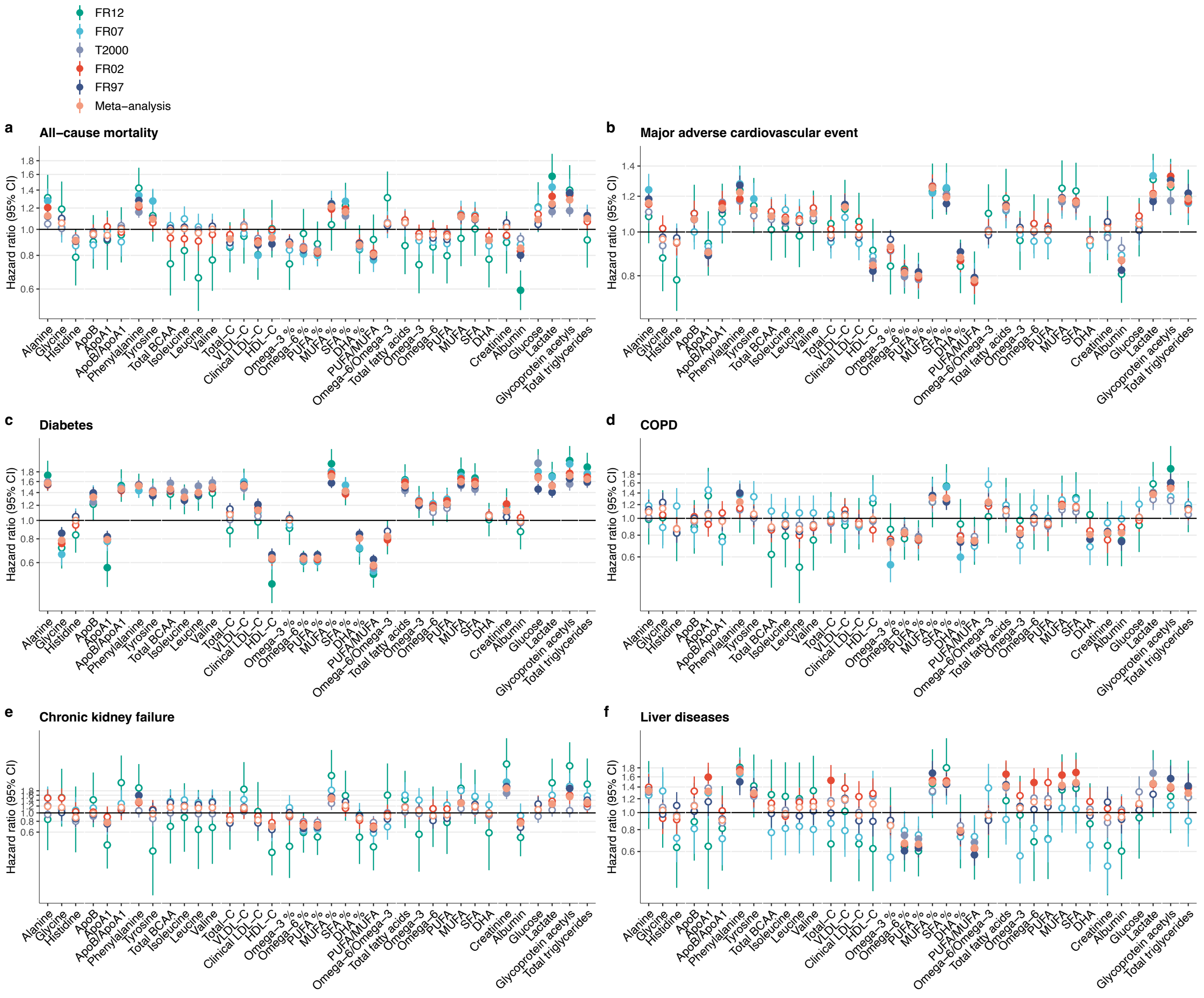


### Supplementary Figure 8. Replication of biomarkers in each cohort.

Replication of the biomarker associations across six endpoints by each cohort in Finnish Institute for Health and Welfare (THL) biobank. Results come from separate analyses of the 5 prospective Finnish cohorts (FINRISK 1997, 2002, 2007, and 2012, and Health 2000) and a meta-analysis of these results. Hazard ratios (HRs) and 95% confidence intervals (CIs) are shown per SD-scaled biomarker concentrations to facilitate comparison across the biomarkers. The black horizontal line denotes a hazard ratio of 1. The biomarkers represent 37 biomarkers that are clinically validated in the Nightingale NMR platform.
