## Appendix for "Atlas of plasma nuclear magnetic resonance biomarkers for health and disease in 118,461 individuals from the UK Biobank"

### Appendix 1

#### Nightingale Health NMR biomarker profiling in the UK Biobank

Nightingale Health Plc. is performing biomarker profiling of baseline plasma samples from all participants in the UK Biobank. Details of the Nightingale Health NMR biomarker platform have been described previously (Soininen et al. 2015; Würtz et al. 2017). Appendix Figure 1 illustrates key steps of the measurement process. The biomarker measurements took place in Finland between 2019 and 2020 using six NMR spectrometers. The first data release covers biomarker measurements from a random selection of approximately 118,481 EDTA plasma samples from the baseline recruitment. In addition, around 4,000 EDTA plasma samples from repeat assessment are included in the same data release, with both baseline and repeat-visit sample measured for ~1,500 participants. The NMR biomarker dataset has been made available for the research community through the UK Biobank in March 2021.

All sample analysis processes were performed according to the standard operating procedures that are part of Nightingale Health's EN ISO 13485 certified Quality Management System (certified by DEKRA Certification B.V.). Nightingale Health measures all blood samples with a CE-marked In Vitro Diagnostic Medical Device and Nightingale Health has received accreditation (SFS-EN ISO/IEC 17025:2005) for its laboratory from the Finnish Accreditation Service FINAS. The scope of accreditation and sites are available at www.finas.fi.

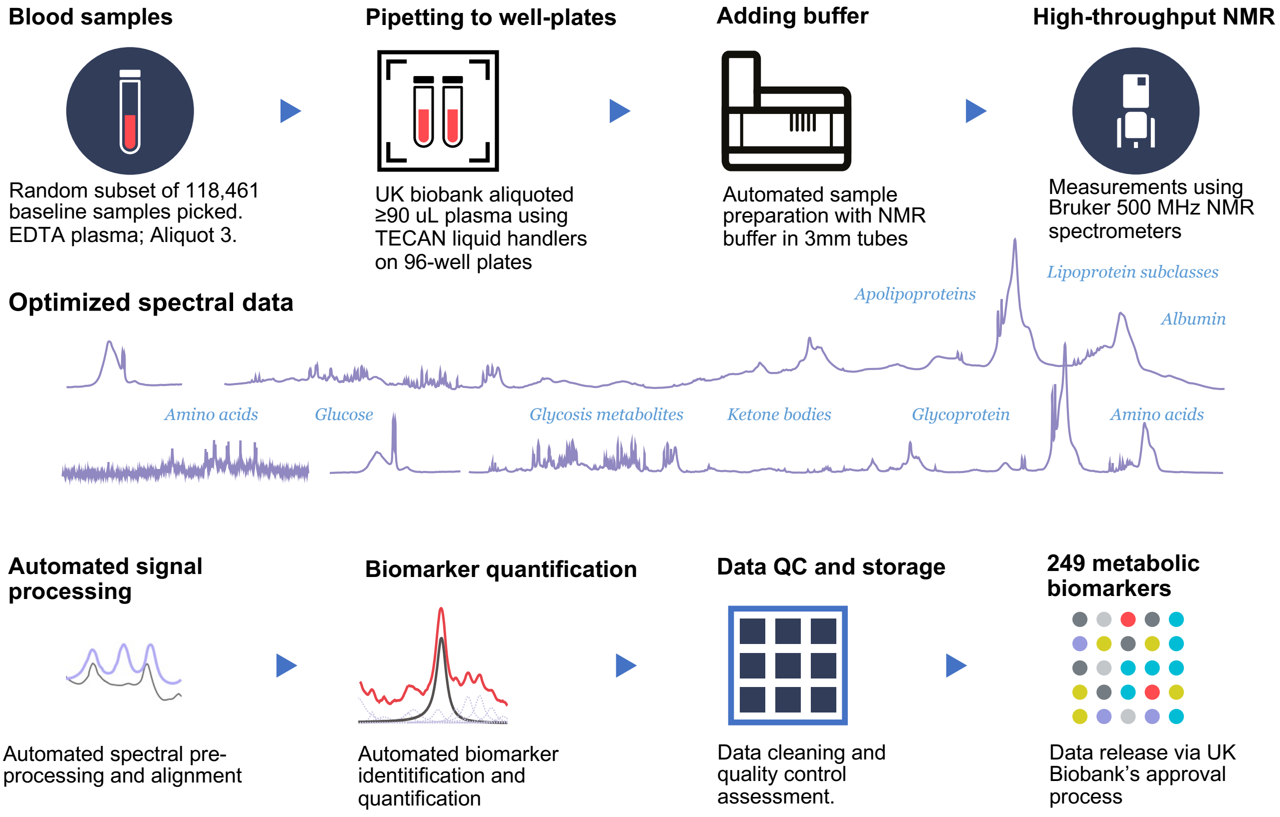


**Appendix Figure 1. Key steps in biomarker measurement process of the Nightingale Health UK Biobank initiative.**

*Plasma sample preparation*

EDTA plasma samples from aliquot 3 were prepared in 96-well plates by UK Biobank laboratory (Stockport, UK). At least 90 $\mu$L plasma was aliquoted in each well using TECAN freedom EVO 150 robotic liquid handlers, which have coefficients of variation (CV) in pipetting volume at <0.75% across 8 tips. The plasma samples were shipped Nightingale Health's laboratories in Finland in the 96-well plates on dry ice in batches of 5,000-20,000 samples. No selection criteria were applied to the sampling, due to convenience of sample picking; the 118,461 samples are therefore a random subset of the full cohort.

Samples were stored in a freezer at -80°C at Nightingale Health laboratories after arrival from UK Biobank laboratory. Before preparation, frozen samples were slowly thawed at +4°C overnight, and then mixed gently and centrifuged (3 min, 3400´*g*, +4°C) to remove possible precipitate. Aliquots of each sample were transferred into 3-mm outer-diameter NMR tubes and mixed in 1:1 ratio with a phosphate buffer (75mM $Na_2HPO_4$ in 80%/20% $H_2O/D_2O$, pH 7.4, including also 0.08% sodium 3-(trimethylsilyl) propionate-2,2,3,3-d4 and 0.04% sodium azide) automatically with an automated liquid handler (PerkinElmer Janus Automated Workstation).

*NMR spectroscopy*

The plasma samples were measured in six NMR spectrometers. Measurements were conducted blinded prior to the linkage to the UK Biobank health outcomes. The prepared plasma samples on 96-well plates were loaded onto a cooled sample changer, which maintains the temperature of samples waiting to be measured at +6°C. Two NMR spectra were recorded for each plasma sample using 500 MHz NMR spectrometers (Bruker AVANCE IIIHD). The first spectrum is a presaturated proton spectrum, which features resonances arising mainly from proteins and lipids within various lipoprotein particles. The other spectrum is a Carr-Purcell-Meiboom-Gill $T_2$-relaxation-filtered spectrum where most of the broad macromolecule and lipoprotein lipid signals are suppressed, leading to enhanced detection of low-molecular-weight metabolites.

*Quantified biomarkers*

The biomarkers were quantified using Nightingale Health's proprietary software (quantification library 2020). This simultaneously quantifies 249 metabolic measures per EDTA plasma sample, comprising 168 in absolute levels and 81 ratio measures (Appendix Figure 2). All the biomarkers are of known-identity. The biomarker panel include routine lipids, lipoprotein subclass profiling with lipid concentrations within 14 subclasses, fatty acid composition, and various low-molecular weight metabolites such as amino acids, ketone bodies and glycolysis metabolites quantified in molar concentration units. For 14 lipoprotein subclasses, the lipid concentrations and composition are measured in terms of triglycerides, phospholipids, total cholesterol, cholesterol esters, and free cholesterol, and total lipid concentration within each subclass. The majority of the biomarkers are measured in absolute concentration units (mmol/L). For clinical translation applications, 37 biomarkers in the panel have been certified for diagnostics use (CE-marked). The average detection rate was >99% across all plasma samples.

**Amino acids (mmol/l)**
* Alanine mmol/l
Glutamine mmol/l
* Glycine mmol/l
* Histidine mmol/l
* Phenylalanine mmol/l
* Tyrosine mmol/l
* Isoleucine mmol/l
* Leucine mmol/l
* Valine mmol/l
* Total branched-chain amino acids

**Glycolysis related metabolites (mmol/l)**
* Glucose
* Lactate
Pyruvate
Citrate

**Ketone bodies (mmol/l)**
3-hydroxybutyrate
Acetoacetate
Acetone
Acetate

**Inflammation (mmol/l)**
* Glycoprotein acetyls (GlycA)

**Fluid balance (mmol/l)**
* Creatinine
* Albumin

**Fatty acids (mmol/l)**
* Total fatty acids
* Omega-3 fatty acids
* Omega-6 fatty acids
* Polyunsaturated fatty acids (PUFA)
* Monounsaturated fatty acids (MUFA)
* Saturated fatty acids
Linoleic acid
* Docosahexaenoic acid (DHA)

**Fatty acid ratios (%)**
* Omega-3 fatty acids ratio to total fatty acids
* Omega-6 fatty acids ratio to total fatty acids
* PUFA ratio to total fatty acids
* MUFA ratio to total fatty acids
* Saturated fatty acids ratio to total fatty acids
Linoleic acid ratio to total fatty acids
* DHA ratio to total fatty acids
* PUFA to MUFA ratio
* Omega-6 fatty acids to omega-3 fatty acids ratio
Degree of unsaturation

**Other lipids (mmol/l)**
Phosphoglycerides
Ratio of triglycerides to phosphoglycerides ratio
Total cholines
Phosphatidylcholines
Sphingomyelins

**Cholesterol (mmol/l)**
* Total cholesterol
Non-HDL-C
Remnant cholesterol
* VLDL cholesterol
* Clinical LDL cholesterol
LDL cholesterol (size-specific)
* HDL cholesterol

**Triglycerides (mmol/l)**
* Total triglycerides
Triglycerides in VLDL
Triglycerides in LDL
Triglycerides in HDL

**Apolipoproteins (g/l)**
* Apolipoprotein B
* Apolipoprotein A1 g/l
* Apolipoprotein B to apolipoprotein A1 ratio

**Lipoprotein particle size (nm)**
Average diameter for VLDL particles
Average diameter for LDL particles
Average diameter for HDL particles

**Lipoprotein particle concentrations (mmol/l)**
Total concentration of lipoprotein particles
Concentration of VLDL particles
Concentration of LDL particles
Concentration of HDL particles

**Total lipids in lipoprotein particles (mmol/l)**
Total lipids in lipoprotein particles
Total lipids in VLDL
Total lipids in LDL
Total lipids in HDL mmol/l

**Phospholipids (mmol/l)**
Total phospholipids
Phospholipids in VLDL
Phospholipids in LDL
Phospholipids in HDL

**Cholesteryl esters (mmol/l)**
Total esterified cholesterol
Cholesteryl esters in VLDL
Cholesteryl esters in LDL
Cholesteryl esters in HDL

**Free cholesterol (mmol/l)**
Total free cholesterol
Free cholesterol in VLDL
Free cholesterol in LDL
Free cholesterol in HDL

**Particle concentration and lipid composition for 14 lipoprotein subclasses**
Particle concentration (mmol/l)
Total lipids (mmol/l)
Phospholipids (mmol/l and % of total lipids)
Cholesterol (mmol/l and % of total lipids)
Cholesteryl esters (mmol/l and % of total lipids)
Free cholesterol (mmol/l and % of total lipids)
Triglycerides (mmol/l and % of total lipids)

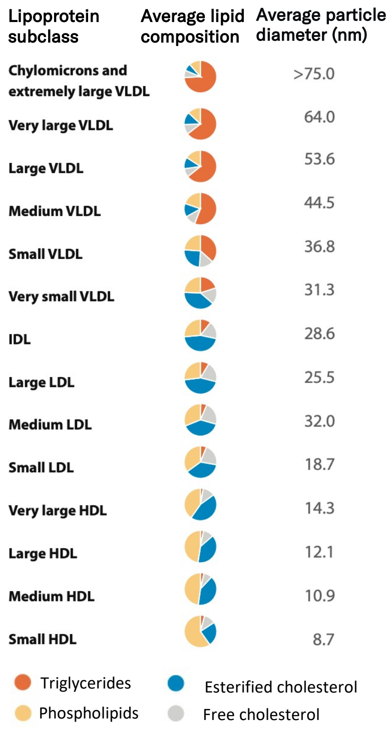


**Appendix Figure 2. Overview of biomarkers quantified by the Nightingale Health NMR platform.** The majority of measures reflect lipid metabolism, but the biomarkers also cover proteolysis, glycolysis, and ketolysis. The biomarkers reflect diverse health aspects such as chronic inflammation, dietary quality and the risk for hundreds of disease endpoints. The 37 biomarkers marked with * are those currently certified for diagnostics use. Lipoprotein subclasses are defined in particle-size specific manner calibrated against gel permeation high-performance liquid chromatography (Würtz & Soininen, 2020). The average particle size of each subclass is indicated. For each of the 14 lipoprotein subclasses 12 measures are provided: the circulating concentration of total lipids in the particles (sum of free and esterified cholesterol, triglycerides and phospholipids), the particle concentration, and the absolute circulating concentration of five main lipids (free, esterified and total cholesterol, triglycerides and phospholipids) and the relative proportions of these five lipids in each particle subclass. The lipoprotein subclasses are defined according to their particle size as illustrated in the figure. HDL indicates high-density lipoprotein; IDL, intermediate-density lipoprotein; LDL, low-density lipoprotein; VLDL, very-low density lipoprotein. Clinical LDL cholesterol and size-specific LDL cholesterol refer to different methods for defining LDL (Holmes & Ala-Korpela 2019).

#### Quality control protocol

Pre-specified metrics on the biomarker consistency were agreed between UK Biobank and Nightingale Health to ensure the quality of results throughout the project. The full project was initiated after the consistency metrics of the pilot were met. Two internal control samples provided by Nightingale Health were included in each 96-well plate for tracking the consistency across multiple spectrometers during the project. Four sets of internal control samples with different biomarker concentration span were used across the 1,352 96-well plates measured. These were interleaved between the NMR instruments for extended periods of the project duration. An example of such continuous quality control is illustrated in Appendix Figure 3 for the case of leucine.

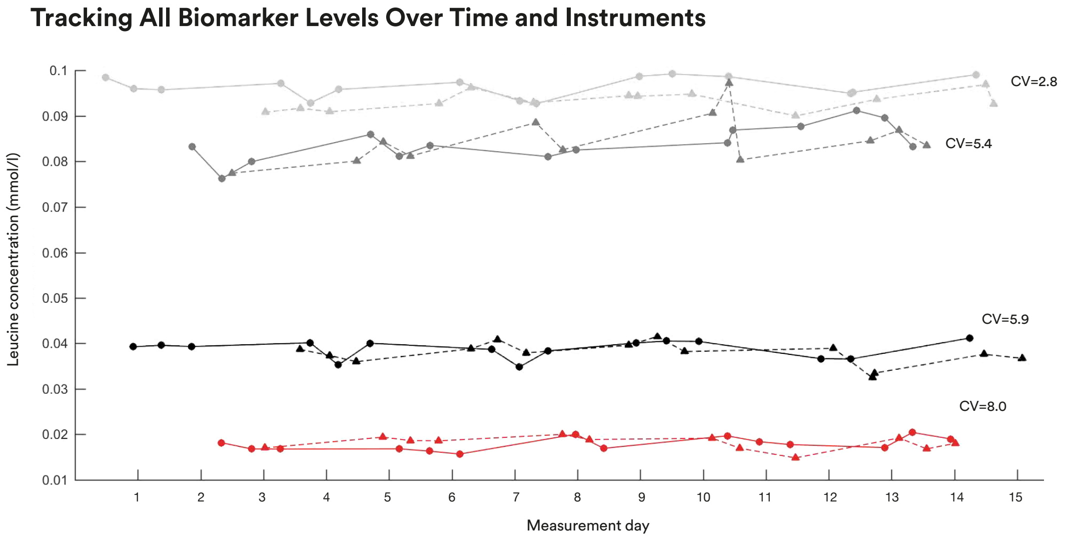


**Appendix Figure 3: Tracking the biomarker quantification along the measurements.** The consistency of the biomarker quantification in the control samples is illustrated for leucine; similar tracking was done for all biomarkers. Different colors show results for four control samples that are measured interleaved in NMR instruments during the project course. Dashed and full-blown lines indicate results from two different NMR instruments.

*Technical and biological repeatability*

Two blind duplicate samples provided by UK Biobank were included on each 96-well plate. The position information of these blind duplicates were unlocked only after interim results delivery to UK Biobank. Appendix figure 4 illustrates the distribution of coefficients of variation (CV) across the biomarker measures, both for the UK Biobank's blind duplicates and Nightingale Health's internal control samples. The CVs are below 5% for the majority of the biomarkers in both instances. CVs of individual biomarkers based on the blind duplicate samples are provided in Appendix Table 1. These results fulfilled the pre-specified CV targets across the biomarker measures for each consecutively measured set of ~20,000 samples. Prior studies on smaller scale have also reported representative CVs for blind duplicate samples as well as for repeat control samples for NMR biomarkers (Holmes et al. 2018; Kettunen et al. 2016).

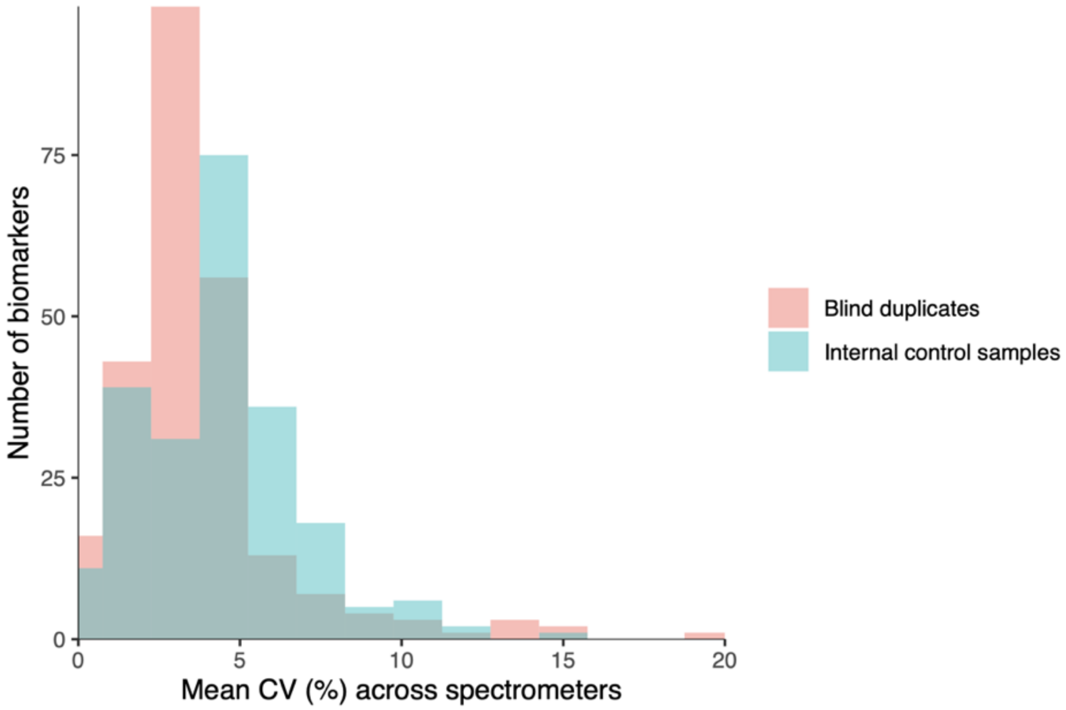


**Appendix Figure 4**. **Distributions of coefficients of variation (CV) for the 249 metabolic measures**. Results for UK Biobank's blind duplicate samples are shown in red and for internal control samples in blue. The CVs are assessed across the six NMR spectrometers used for the measurements. CVs of individual biomarkers are available in Appendix Table 1.

The technical consistency of measurements over consecutive shipment batches and in different NMR spectrometers are illustrated for all the NMR biomarkers in the UK Biobank data resource (https://biobank.ndph.ox.ac.uk/showcase/label.cgi?id=220). This resource also shows the correlation for blinded duplicate samples for each biomarker, as well as the biological consistency in repeat-visit samples drawn from the same individuals four years apart. These assessments of the technical and biological repeatability are illustrated for GlycA in Appendix Figure 5.

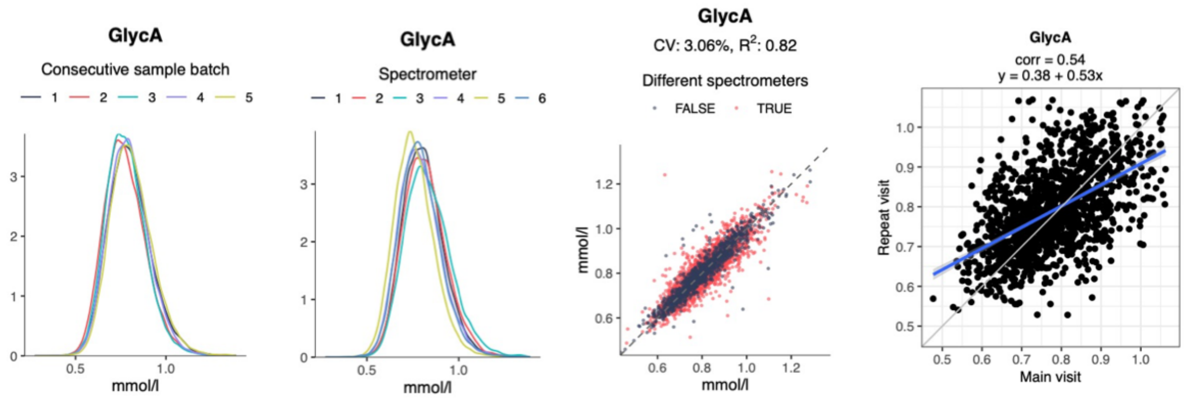


**Appendix Figure 5**. **Technical and biological repeatability for glycoprotein acetyls (GlycA).** The technical consistency over time is shown in terms of distributions of consecutive batches of sample shipments (first panel) and in different spectrometers (second panel). The technical repeatability is also shown in form of the consistency of ~650 blind duplicates samples (third panel), giving rise to an between-instrument CV of 3%. The biological repeatability for measurements from blood samples from the same individuals drawn ~4 years apart is shown for ~1300 samples (forth panel).

*Quality control flags*

The Nightingale Health NMR platform involves integrated quality procedures to reports signs of degradation and contamination issues in each plasma sample. These are reported as flags along with the biomarker concentration result data. Issues affecting the whole sample are reported as sample-level flags; issues affecting only certain biomarkers are reported as biomarker-level flags, provided as a separate data field for each biomarker. In general, if a biomarker has a flag but the value is still provided, it indicates that the presence of the interfering substance is low and deemed not to interfere with the quantification of the biomarker (i.e., the value can be trusted). There is no need to a priori remove any biomarker values based on the flags; however, researchers may consider performing sensitivity analyses as described in the section "Recommended approaches for data pre-processing and epidemiological analyses".

**Plasma sample issues**

*Dilution*

All UK Biobank blood samples are known to suffer from unintended dilution during the initial sample storage process at UK Biobank facilities. Prior reports have suggested that samples from the aliquot 3, used for the NMR measurements, suffer from 5-10% dilution (Allen et al. 2020). The dilution is believed to come from mixing of participant sample with water due to seals that failed to hold a system vacuum in the automated liquid handling systems. While this issue is likely to have an impact on some of the absolute biomarker concentration values, it is expected to have limited impact on epidemiological analyses. However, we recommend that this aspect is considered when conducting analyses that rely on absolute concentrations, such as stratification based on biomarker quantification. This may also cause challenges to compare distributions of biomarker concentrations to those observed in other cohort studies, and we therefore caution against using the concentrations observed in UK Biobank as reference levels for translational applications.

*Technical variation and outlier plates*

An independent study from Cambridge University conducted post-measurement quality control and analysed sources of technical variation in the NMR biomarker data (Ritchie et al. 2021). In short, this study identified spectrometer used and time from plasma preparation to measurement as the only two notable sources of variation. Each factor explained 1-3% of variation for the majority number of biomarkers, and only amino acids histidine and alanine had substantially higher for technical variation. Researchers may consider regressing out or adjusting for spectrometer as a factor in epidemiological analyses. An R package has been made available to remove such technical variation. This can especially provide slight power gains for genome-wide association analyses, whereas the impact on biomarker-disease associations is minor.

The same study highlighted a small number of outlier plates with deviating concentration across many biomarkers (Ritchie et al 2021). This technical variation was deemed to arise from UK Biobank's sample plating process. A median of 9 outliers plates were identified across the biomarkers, with a maximum of 20 for albumin. The authors recommended to remove data from these outlier plates; however, with only ~1% of the samples affected the impact on epidemiological associations is modest.

#### Comparison to clinical chemistry and mass spectrometry

##### *Comparison to serum biochemistry in UK biobank and THL Biobank*

If comparisons are done to the clinical biochemistry biomarkers available in the UK Biobank, it is important to note that those were measured from serum samples, primarily from aliquot 1. In contrast, the NMR-based biomarkers were measured from EDTA plasma samples from aliquot 3. Different aliquots are affected by different degree of dilution (Allen et al. 2020). Appendix Figure 6 shows scatterplots and correlation coefficients between lipids, apolipoproteins, creatinine, albumin and glucose measured both by routine clinical chemistry assays and NMR in the UK Biobank. For comparison, we also show corresponding plots showing the biomarker consistency in the FinHealth 2017 study, a population-based cohort under the Finnish Institute for Health and Welfare (THL) Biobank (n~6,000) (Borodulin et al., 2017). In the FinHealth 2017 cohort, clinical chemistry assays were measured in a central laboratory from frozen serum samples soon after the cohort survey and the NMR biomarkers were measured 1-2 years after from frozen serum samples during 2018-19 using 350 $\mu$L aliquots of serum.

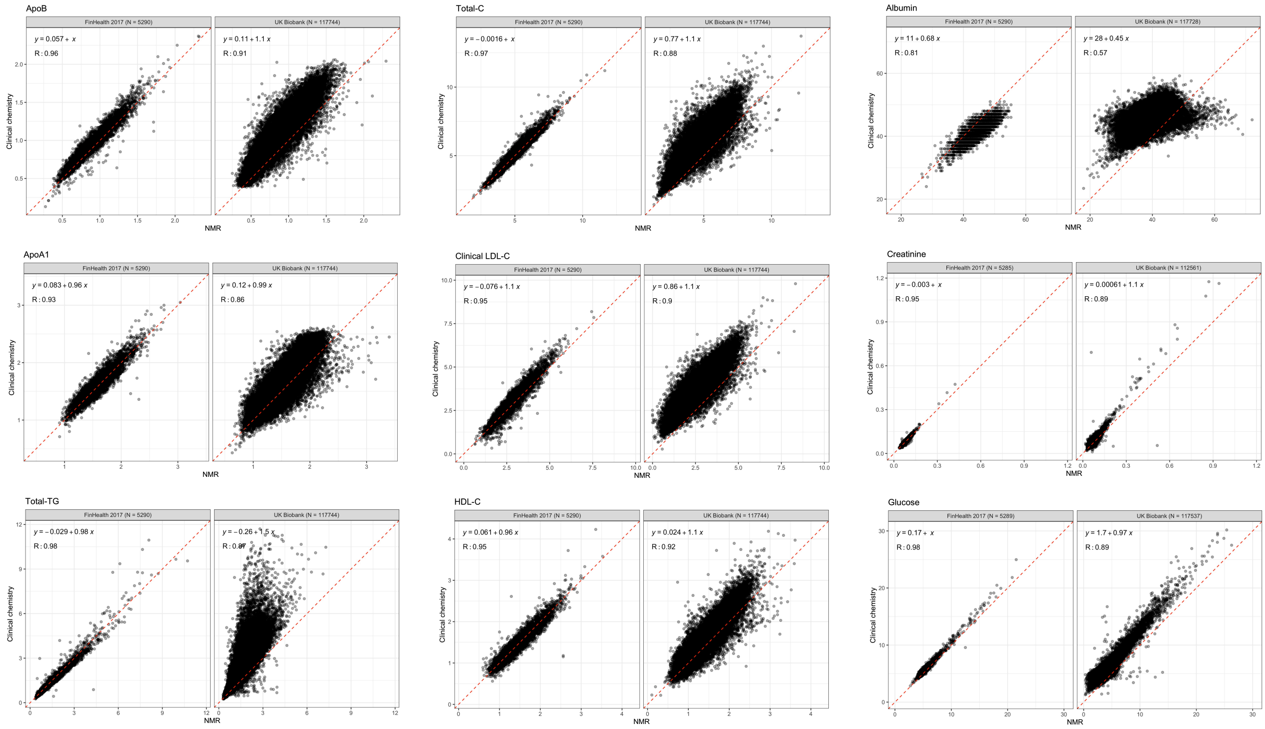

**Appendix Figure 6**. Scatter plots of lipids and other routine biomarkers quantified with NMR platform and clinical chemistry analyzers for the UK Biobank (n=118,000) and the FinHealth 2017 cohort (n~6,000). Correlation coefficients (R) represent linear Pearson's correlations. The regression line and the corresponding equation represent the slope and offset from ordinary least squares linear regression fit. Clinical biochemistry was measured using Beckman Coulter AU5800 instruments, with direct LDL-C measured by enzymatic selective protection.

Correlation coefficients were high in both cohorts, but the overall consistency was weaker in UK Biobank compared to the FinHealth 2017 cohort. In addition, the offset and slope deviations from the diagonal were more pronounced in UK Biobank. This is likely owing both to the sample dilution issue where different aliquots used for the measurements, as well as between serum and EDTA plasma and differences in sample storage time. The correlation coefficient for albumin in particular was weak in the UK Biobank; however, we note that the associations with disease outcomes were broadly similar for albumin for both assays.

The consistency of the NMR biomarker measurements with clinical biochemistry are in line with earlier studies that have reported correlation coefficients above 0.9 (Würtz et al. 2017). A recent study reported correlations of the same NMR biomarkers with clinical chemistry for FINRISK cohorts under the THL Biobank: the correlations were r~0.95 for the newest sample collection (FINRISK 2012), and r~0.90 for the oldest sample collections (FINRISK 1997). (Tikkanen et al. 2021).

*Comparison with other multi-biomarker platforms*

The biomarker coverage from the Nightingale Health NMR platform is mostly distinct from those of mass-spectrometry based assays (Würtz et al. 2017). Less than 20 out the 249 biomarkers are quantified by the main mass-spectrometry metabolomics vendors. Only in the case of amino acids and glycolysis metabolites is there direct overlap with mass spectrometry platforms. The main reason for the limited biomarkers overlap is that mass-spectrometry platforms are generally not able to quantify the detailed lipoprotein measures obtained by the Nightingale Health NMR platform, since the physiological character of lipoprotein particles are destroyed in mass spectrometry. Furthermore, important analytes not commonly analyzed by mass spectrometry include the GlycA composite-protein biomarker as well as aggregate fatty acid measures (e.g. omega-3%), which are relevant for dietary studies and supplementation trials and often more interpretable than molecule-specific fatty acids.

The measurements of fatty acids, amino acids and glycolysis metabolites by the Nightingale Health NMR platform have been certified for clinical use. To further demonstrate the validity of NMR biomarker quantification, we here report previously unpublished correlations of amino acids, glycolysis metabolites and circulating fatty acids measured with three other analytical platforms. We note that these measurements were not done at the same time from split aliquots, and do therefore not represent strict analytical comparisons but rather consistency in cohort settings, potentially from different blood specimens and measured years apart.

Appendix Figure 7 shows the scatter plots of absolute and relative fatty acids measured by Nightingale Health NMR platform in comparison to gas chromatography (Vitas Analytical Services, Oslo, Norway). The samples are from a familial hypercholesterolemia cohort of N=144 individuals (Øyri et al. 2018). The correlations were especially high for absolute fatty acid measures (r=0.89-0.98) and slightly lower for fatty acid ratios, relative to total fatty acids (r=0.80-0.92). These results are consistent with comparisons of NMR vs gas chromatography fatty acids, using a prior version of the Nightingale Health biomarker platform involving a lipid extraction step (Würtz et al. 2017).

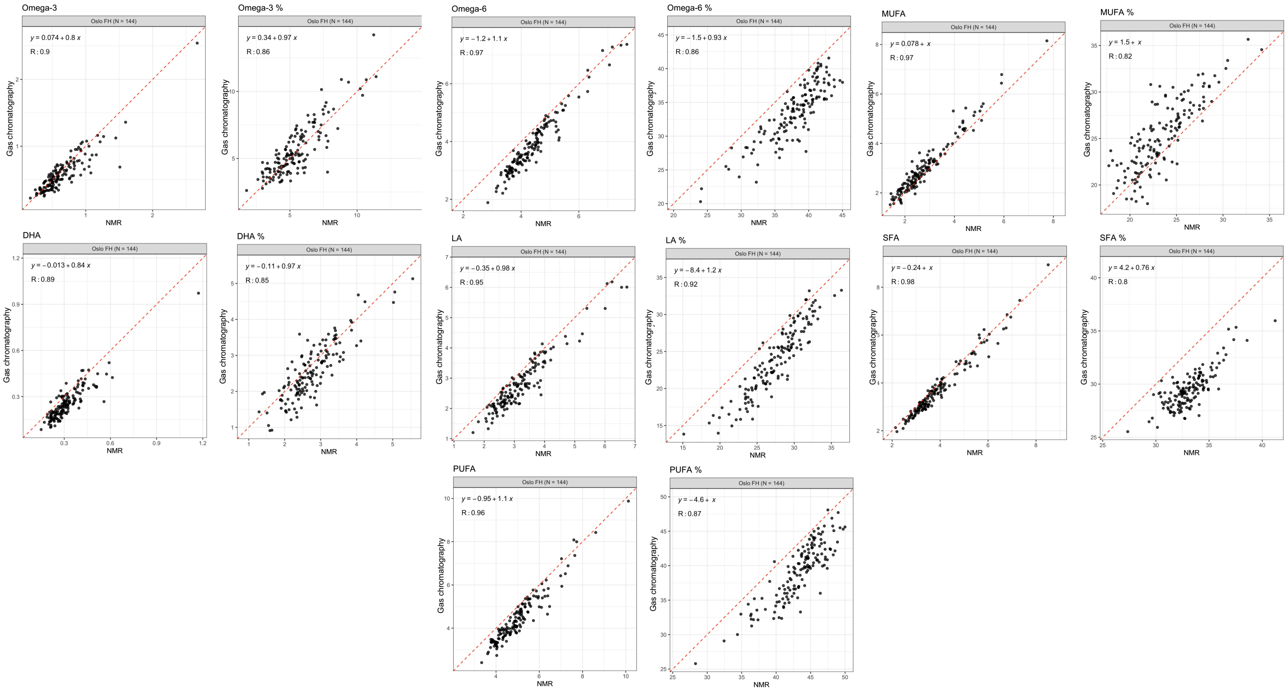


**Appendix Figure 7**. Scatter plots of fatty acids quantified with NMR and gas chromatography (n=144; study on familial hypercholesterolemia from University of Oslo, Norway; Øyri et al, 2018). Correlation coefficients (R) represent linear Pearson's correlations. The regression line and the corresponding equation represent the slope and offset from ordinary least squares linear regression fit.

Appendix Figure 8 shows the scatter plots of amino acids in comparison to the Biocrates p180 mass spectrometry platform (Innsbruck, Austria) in the ADNI1 cohort (N=749). Correlations were highest for branched-chain and aromatic amino acids as well as alanine and glycine (r=0.78-0.90) and lower for glutamine (r=0.65) and histidine (r=0.54).

Appendix Figure 8 also shows the scatter plot for the ketone body 3-hydroxybutyrate in comparison to measured to a cyclic enzymatic method (Wako Chemicals GmbH, Neuss, Germany) in an Italian cohort (Tikkanen et al. 2019). The correlation with NMR-based measure was r=0.98. From the same study, the triglyceride-rich lipoprotein cholesterol measured by the Nightingale Health NMR platform has previously been reported to correlate well with ultra-centrifugation (r=0.90; Würtz & Soininen 2020).

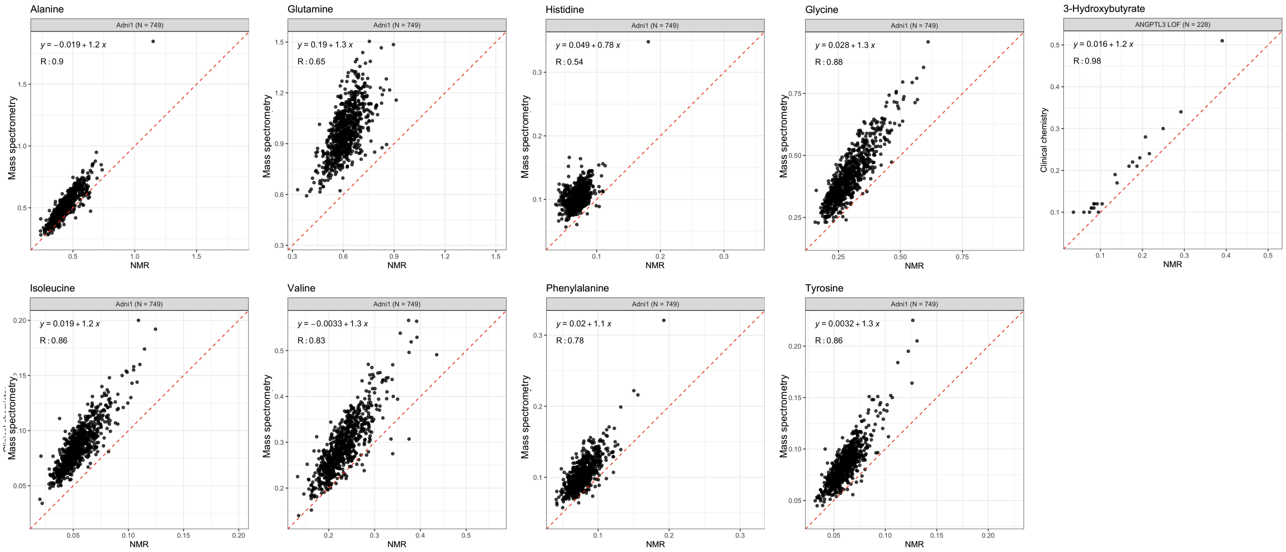


**Appendix Figure 8**. Scatter plots of amino acids quantified with Nightingale Health NMR platform in comparison to Biocrates p180 mass spectrometry in the ADNI1 cohort (n=749). The right most plot shows the comparison of 3-hydroxybutyrate measured with an enzymatic method in an Italian study of postprandial effects in ANGPTL3 loss-of-function carriers and their controls (n=228; Tikkanen et al 2019). Correlation coefficients (R) represent linear Pearson's correlations. The regression line and the corresponding equation represent the slope and offset from ordinary least squares linear regression fit.

Finally, Pearson's correlations of amino acids and glycolysis-related metabolites measured by the Nightingale Health platform and the Metabolon HD4 mass-spectrometry platform (Morrisville, North Carolina, US) from the same samples were the following in the Qatar Metabolomics Study on Diabetes cohort (QMDiab): leucine r=0.86; valine r=0.82; phenylalanine r=0.67; tyrosine r=0.90; glutamine r=0.75; histidine r=0.62; alanine r=0.75; glucose r=0.86; lactate r=0.93; citrate r=0.81 (results courtesy of Karsten Suhre, Weill Cornell Medicine Qatar). These results are consistent with other studies reporting the medium to high consistency (r=0.42-0.85) of the few biomarkers overlapping between Nightingale Health NMR and mass-spectrometry data from Metabolon and Biocrates (Deelen et al. 2019; Schmidt et al. Metabolites 2021).

**Recommended data processing for epidemiological analyses**

The NMR biomarker data in UK Biobank can generally be used for epidemiological analyses without any pre-processing, and can as a rule of thumb be analysed in the same manners as the clinical biochemistry data available in UK Biobank. The clinical chemistry data, already available in the full cohort, can also be used as positive control in case of overlapping measures, and a means to put association magnitudes into context of established clinical measures. The approach of 4SD outlier exclusion and log-transformation chosen for the biomarker-disease atlas (biomarker-atlas.nightingale.cloud/)) is applied to have a consistent approach, but omission or variations of these steps generally has minute influence on the biomarker-disease associations.

The degree of missingness of any biomarker is generally small (<1%). For analyses of samples marked as zero concentration value we can recommend replacing the zero values with the values just below the lowest observed value, since it avoids artificial drops in the distributions. For analyses that require complete data of multiple or all biomarkers simultaneously, we recommend methods for imputation rather than excluding the entire sample if there is a few missing biomarkers.

*Accounting for quality control flags*

Biomarker values substantially affected by interfering substances have been removed during the quality control procedures. However, the researcher may consider performing sensitivity analyses by excluding samples flagged with “Low protein”, which may indicate more severe sample dilution. Biomarker values flagged with “Below limit of quantification” may also be omitted in sensitivity analyses, since this flag indicates that the concentration of the given biomarker is smaller than the range where the quantification of the NMR biomarkers is considered highly accurate. The analyses done for the biomarker disease-atlas does not omit these values.
